# The SARS-CoV2 envelope is distinct from host membranes, exposes pro-coagulant lipids, and can be inactivated *in vivo* by surfactant-containing oral rinses

**DOI:** 10.1101/2022.02.16.22270842

**Authors:** Zack Saud, Victoria J Tyrrell, Andreas Zaragkoulias, Majd B Protty, Evelina Statkute, Anzelika Rubina, Kirsten Bentley, Daniel A. White, Patricia Dos Santos Rodrigues, Robert C Murphy, Harald Köfeler, William J Griffiths, Jorge Alvarez-Jarreta, Richard William Brown, Robert G Newcombe, James Heyman, Manon Pritchard, Robert WJ Mcleod, Arvind Arya, Ceri-Ann Lynch, David Owens, P Vince Jenkins, Niklaas J. Buurma, Valerie B O’Donnell, David W. Thomas, Richard J. Stanton

## Abstract

The lipid envelope of SARS-CoV2 is an essential component of the virus, however its molecular composition is unknown. Addressing this knowledge gap could support the design of anti-viral agents, and further understanding of viral interaction with extracellular host proteins, infectivity, pathogenicity, and innate immune system clearance. Lipidomics analysis of SARS-CoV2 particles generated from Vero or A549 cells revealed that the virus envelope comprised mainly of phospholipids (PL), primarily phosphatidylcholine (PC), phosphatidylethanolamine (PE) and phosphatidylinositol (PI), with very little cholesterol, sphingolipids or other lipids, indicating significant differences from host membranes. Unlike healthy cellular membranes, procoagulant aminoPL (aPL), specifically PE and phosphatidylserine (PS), were present on the external side at levels far exceeding those seen on activated platelets. As a result, purified virions directly promoted coagulation. To investigate whether these differences enabled the viral envelope to be selectively targeted at relevant sites *in vivo*, we tested whether non-toxic oral rinses containing lipid disrupting chemicals could reduce viral infectivity. Products containing PL-disrupting surfactant solutions (cetylpyridinium chloride (CPC) or ethyl lauroyl arginate) met EN14476 virucidal standards *in vitro*, however products containing essential oils, PVP-I, or Chlorhexidine did not, nor did rinses containing components that altered the critical micelle concentration of CPC. This result was recapitulated *in vivo*, where a 30-second oral rinse with CPC-mouthwash eliminated live virus in the oral cavity of COVID19 patients for at least 1hr, while PVP-Iodine and saline mouthwashes were ineffective. Thus, the SARS-CoV2 lipid envelope is distinct from the host plasma membrane which may enable design of selective anti-viral approaches, it exposes PE and PS which may influence thrombosis, pathogenicity, and inflammation, and can be selectively targeted *in vivo* by specific oral rinses.

## Introduction

The lipid envelope is critical to the structure and function of SARS-CoV2, as for all enveloped viruses, such as influenza, HIV, herpes simplex virus, MERS and SARS-CoV (*1, 2*). Yet despite this, the potential of the envelope as an antiviral target has not been exploited, beyond being the target of handwashing and gels, where soap or high concentrations of ethanol (>60%) dissolve the lipids and inactivate the virus. This is in part because, unlike our extensive knowledge of the structure and function of the proteins in the virion (*1, 2*), there is no information on the lipid composition of the SARS-CoV2 envelope – indeed, viral lipid envelopes overall are surprisingly unstudied, and their detailed lipid composition unknown.

Coronaviruses bud from the endoplasmic reticulum/Golgi intermediate complex (ERGIC) and exit via lysosomal secretion (*3-8*), thus the composition of the virion envelope may differ significantly from plasma membrane, enabling selective therapeutic targeting that avoids damaging host membranes (*9*). Furthermore, the envelope is not simply a structural component of the virion, with lipids themselves being potent bioactive molecules. Mammalian cells maintain aminophospholipids (aPL), such as phosphatidylethanolamine (PE) and phosphatidylserine (PS), in their inner plasma membrane leaflet using energy-dependent enzymes, however these control mechanisms are not present in the virus. This raises the possibility that the external face is enriched in PE and PS, which are highly pro-thrombotic, and furthermore could directly promote virion uptake via apoptotic cell mimicry (*10-16*). Indeed, a recent study showed that PS is present on the surface of the virions and that PS receptors on host cells can support entry (*17*). However, that study relied on an ELISA method and neither the amounts nor the molecular species of PS exposed were demonstrated, nor was the presence of PE shown. Phospholipids (PL) such as lysophospholipids and sphingolipids/ceramides are pro-inflammatory effectors (*18, 19*), and can interact with complement to promote a pro-inflammatory environment (*3, 11, 20*), while lysophospholipids signal through G-protein coupled receptors causing immune cell migration and apoptosis (*21-24*). Understanding virion lipid composition therefore has potential to inform our understanding of virus pathogenesis, dissemination, and how the virion promotes transition from early infection to severe inflammatory thrombotic COVID19.

Following on from public health advice on handwashing, which disrupts the lipid envelope, we considered whether similar approaches using formulations that are non-toxic *in vivo* could represent potential anti-viral strategies directed at reducing SARS-CoV2 transmission, and published an evidence review on this topic in 2020(*25*). The lipid membranes of enveloped viruses, including some coronaviruses had previously been shown to be sensitive to disruption by lipidomimetic agents and surfactants(*25*). Thus, we hypothesised that the SARS-CoV2 virus might also be susceptible to inactivation by components in widely-available oral rinses, such as ethanol/essential oils, cetylpyridinium chloride (CPC) and povidone-iodine (PVP-I)(*25*). If lipid-disrupting components in oral rinses can dissolve the virion envelope, this approach could in theory reduce the risk to healthcare workers or carers treating individuals asymptomatically (or symptomatically) carrying the virus. Early in the pandemic, mouthwashes were employed empirically in outbreaks in China but without evidence of efficacy (*26*). Since then, a series of studies have emerged indicating that some can inactivate SARS-CoV2 *in vitro*, including a systematic review (*27-32*). Furthermore, a recent small study suggested that oral rinsing could shorten hospital stay, while another suggested that oral and nasal rinsing could reduce both disease and symptoms in healthcare professionals (*33, 34*). Recently, WHO included a recommendation that PVP-I could be used to reduce the risk of clinical transmission in dentistry (https://www.who.int/publications/i/item/who-2019-nCoV-oral-health-2020.1). However, despite all these encouraging studies, the relative efficacy and the persistence of mouthwashes *in vivo* is currently unknown. Importantly, in order to most effectively target the virus in the oropharynx, a detailed knowledge of the lipid composition is required, so that the most appropriate formulation is selected.

To address these questions, we used lipidomics to provide the amount and molecular diversity of envelope lipids and the levels of external facing aPL in virus cultured from two different cell lines. Our data provides the first complete characterisation of a viral lipid envelope and shows a PL rich membrane that also contains several lysoPL, but is relatively low in cholesterol, sphingomyelin (SM) and other lipids. Sufficient aPL were present to enhance coagulation of plasma in vitro using live virus. Following this, *in vitro* studies tested the interaction of varying lipid-membrane disrupting mouthwash formulations and components. Importantly, only a subset of rinses demonstrated efficacy, specifically those containing surfactant- and polar components. Furthermore, a randomised controlled clinical study in COVID 19 patients showed the virucidal effect of a surfactant-containing rinse against SARS-CoV2 in hospitalised patients. These studies demonstrate the accessibility and importance of lipids as a potential target for anti-viral approaches, which is unlikely to be impacted by mutation of the virus. They also suggest that targeting virus lipids in the oropharynx may be an important component of risk management in healthcare during the COVID19 pandemic, and in the context of other enveloped respiratory viruses including coronaviruses and seasonal influenza viruses in the future.

## Results

### The SARS-CoV2 envelope from Vero or A549 cells is phospholipid (PL) rich and varies depending on cellular origin

SARS-CoV2 England 2 strain was grown in VeroE6 or A549 monolayers and purified using density gradient centrifugation. A549 stably expressed ACE2 and TMPRSS2 to enhance infectivity (*35*). Purity of virus was confirmed by Nanoparticle tracking analysis, with a single peak observed at approximately 100nm (Figure 1 A), and by western blot demonstrating absence of actin as a cellular marker from the purified virus (Figure 1 B, Supplementary Figure 1 A,B). The absence of serum lipid contamination in purified virus was confirmed using precursor scanning for PE, PC and CE (data not shown).

**Figure 1.**
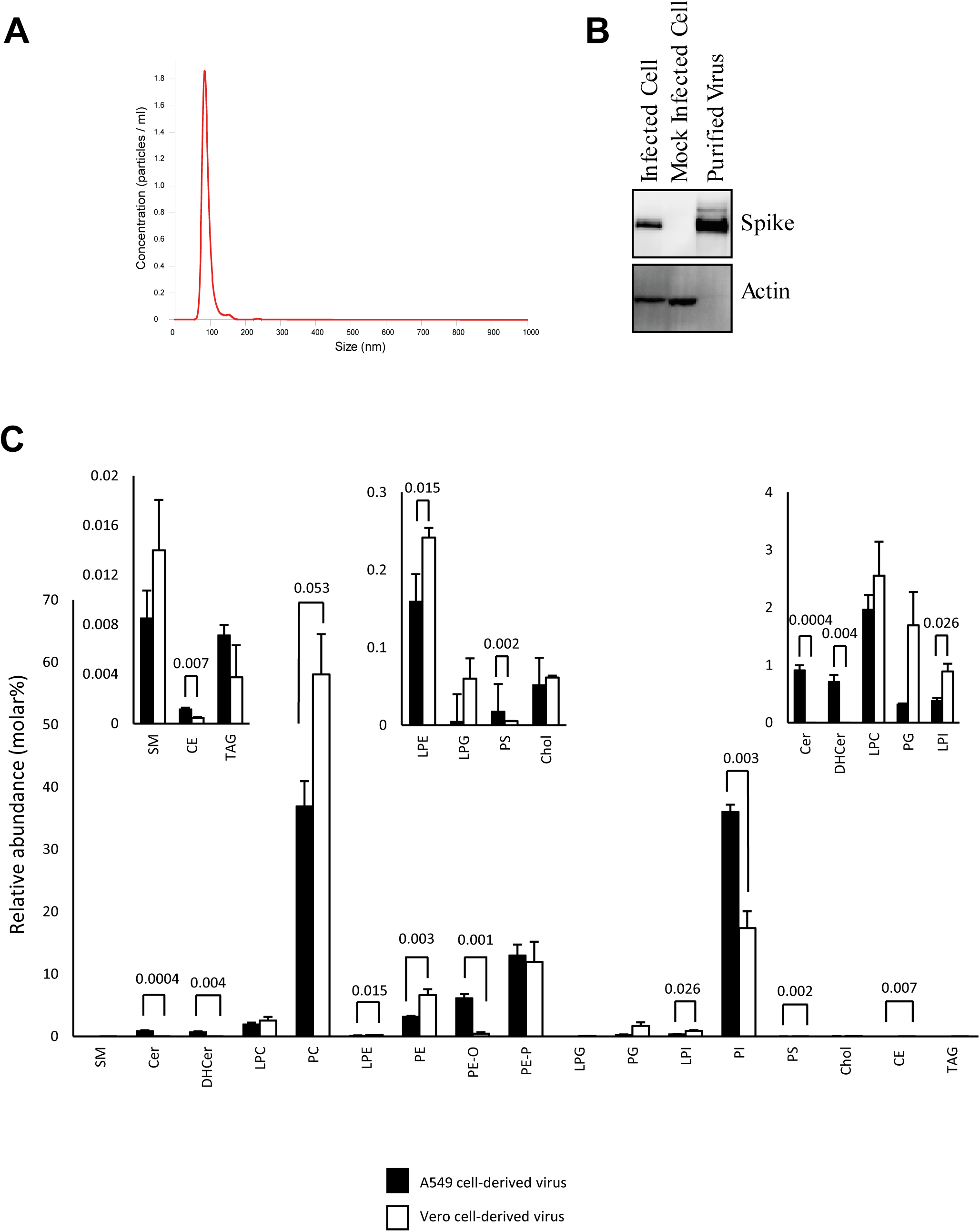
Nanoparticle tracking analysis and western blotting confirm purity of SARS-CoV2 preparations, while targeted lipidomics demonstrates the membrane as a PL rich membrane mainly comprised of PE, PC and PI. *Panel A-B purity analysis of gradient purified viral preparations*. (A) Gradient purified virus was analysed by nanoparticle tracking analysis and particle size plotted. (B) Proteins was solubilised in NuPAGE LDS buffer then separated by size on Bis-Tris gels, before being transferred to nitrocellulose and blotted for the indicated proteins, as outlined in Methods (see Supplementary Figure 1 for uncropped gels) *Panel C. Lipidomics analysis of the total amounts of lipids in each category in Vero and A549 cells*. Lipids were extracted from 3 preparations of virus from either Vero or A549 cells, and analysed using LC/MS/MS, as indicated in Methods. The relative % of all detected lipid categories for all 3 preparations, with molecular species within each category combined to provide total values are shown. Amounts (ng) of all individual molecular species were added together for each preparation then converted to molar amounts using an average mass value per category. Molar% was then calculated following totalling of all lipid categories (n = 3, mean +/- SEM). Unpaired Students T test.

Viral lipid extracts were analysed using lipidomics, including targeted (virus from both cell types) and untargeted (Vero cell virus only) to provide a comprehensive map of molecular composition and abundance. First, targeted LC/MS/MS was used to analyse ∼500 individual molecular species, in triplicate for each cell type. Across the two separate preparations (Vero vs A549), ∼260 lipids were reproducibly detected. The full list of species analysed, and the dataset is provided in Supplementary Data. These data are first shown with lipids grouped into their respective categories (Figure 1 C). Here, data were converted to relative abundance in mol%. This was calculated using a generic mass value for each category of a typical molecular species. Overall, the virus envelope was primarily comprised of PL from several categories, with the most abundant for both preparations being phosphatidylcholine (PC), phosphatidylethanol (PE) and phosphatidylinositol (PI), along with their respective lyso, and ether/plasmalogen forms. Ether/plasmalogen PEs were relatively abundant when compared with acylPEs. Smaller amounts of phosphatidylserine (PS) and phosphatidylglycerol (PG) were seen. There was a low abundance of other lipids such as sphingolipids, including sphingomyelin (SM), ceramide (Cer), dihydroceramides (DHCer), and also cholesteryl esters (CE), triacylglycerides (TAG) and free cholesterol (Figure 1 C). This pattern overall was quite consistent across both virus preparations. However, looking in more detail, some clear differences were also apparent, depending on cell of origin. Comparing A549 with Vero-derived virus, a higher proportion of PI versus PC was seen, along with a higher ratio of etherPE (PE-O), but lower PC and diacylPE (Figure 1C). Some significant differences in low abundance lipid categories such as Cer/DHCer, LPI, LPE, PS and TAGs were also seen (Figure 1 C insets).

### The SARS-CoV2 membrane contains low amounts of cholesterol, SM and PS, relative to other PL

Next, mol% was calculated for PL and sphingolipids only, since this allows comparison with older studies on composition of intracellular membranes of mammalian cells, which used thin layer chromatography coupled with phosphate analysis to measure these lipid categories (Table 1). Unfortunately, very few studies on cell membrane composition exist and these used older methods very different to LC/MS/MS, as well as very different cell types. Nonetheless, it is useful to compare these with SARS-CoV2, as shown in Table 1 (see Discussion). For both virus preparations, the molar ratio of cholesterol:PL was similar, at 0.0005 or 0.00061 mol:mol, A549 or Vero, respectively. This indicates that the membrane is virtually devoid of cholesterol, in combination with a high PL content. Additionally, the mol% of SM and PS are relatively low (Table 1). Overall, the data characterises SARS-CoV2 as a membrane highly enriched in PL, primarily PE, PC and PI.

**Table 1.** Phospholipid composition of SARS-CoV2, compared with rat liver membranes, reproduced from Vance (*45*) and Van Meer (*9*). Approximate phospholipid content is given as % total lipid phosphorus, data is averaged from several sources in both studies, as described.

| Vance (45) |  |  |  |  |  |  |  |  |  |
| --- | --- | --- | --- | --- | --- | --- | --- | --- | --- |
| Lipid | ER | Mito inner | Mito outer | Lysosomes | Nuclei | Golgi | Plasma membrane | Vero | A549 |
| PC | 57 | 41 | 49 | 42 | 52 | 45 | 43 | 61 | 40 |
| PE | 21 | 38 | 34 | 21 | 25 | 17 | 21 | 19 | 23 |
| SM | 4 | 2 | 2 | 16 | 6 | 12 | 23 | 0.024 | 0.009 |
| PI | 9 | 2 | 9 | 6 | 4 | 9 | 7 | 18 | 37 |
| PS | 4 | 1 | 1 | 1 | 6 | 4 | 4 | 0.013 | 0.005 |
| CL | 0 | 16 | 5 | 0 | 0 | 0 | 0 |  |  |
| Other | 5 | <1 | <1 | 14 | 7 | 13 | 2 | 1.75 | 0.33 |
| Chol/PL molar ratio | 0.07 | 0.06 | 0.06 | 0.49 |  | 0.15 | 0.76 | 0.0006 | 0.0005 |
| Van Meer (9) |  |  |  |  |  |  |  |  |  |
| PC | 58 |  |  |  |  | 50 | 39 |  |  |
| PE | 22 |  |  |  |  | 20 | 23 |  |  |
| SM | 3 |  |  |  |  | 8 | 16 |  |  |
| PI | 10 |  |  |  |  | 12 | 8 |  |  |
| PS | 3 |  |  |  |  | 6 | 9 |  |  |
| Chol/PL molar ratio | 0.08 |  |  |  |  | 0.16 | 0.35 |  |  |

Next the individual molecular species within categories were compared. Within each cell type, the levels of specific lipids were very similar, indicating that the viral lipidome is relatively stable (Figure 2, 3A). However, significant differences were seen between Vero *versus* A549-derived virus across many lipids, when comparing fatty acyl (FA) composition (Figure 2,3A). Consistently, levels of PL with the low abundant FA 20:1 and 20:2 were more predominant in Vero than A549 cells, across PE, PG and PI lipids. The pattern was reversed for 20:3 and 20:4, which were more abundant in A549 cells for PE and PI species of PL (Figure 3 B). Aside from this, for more abundant PL species, the pattern was variable with some higher in Vero and others higher in A549 cells (Figure 2, 3A). Notably, for both virus preparations, the most abundant FAs detected were 16:0, 18:0, and 18:1 (Figure 2, 3A). This profile was maintained strongly across all PL classes, as well as lysoPLs.

**Figure 2.**
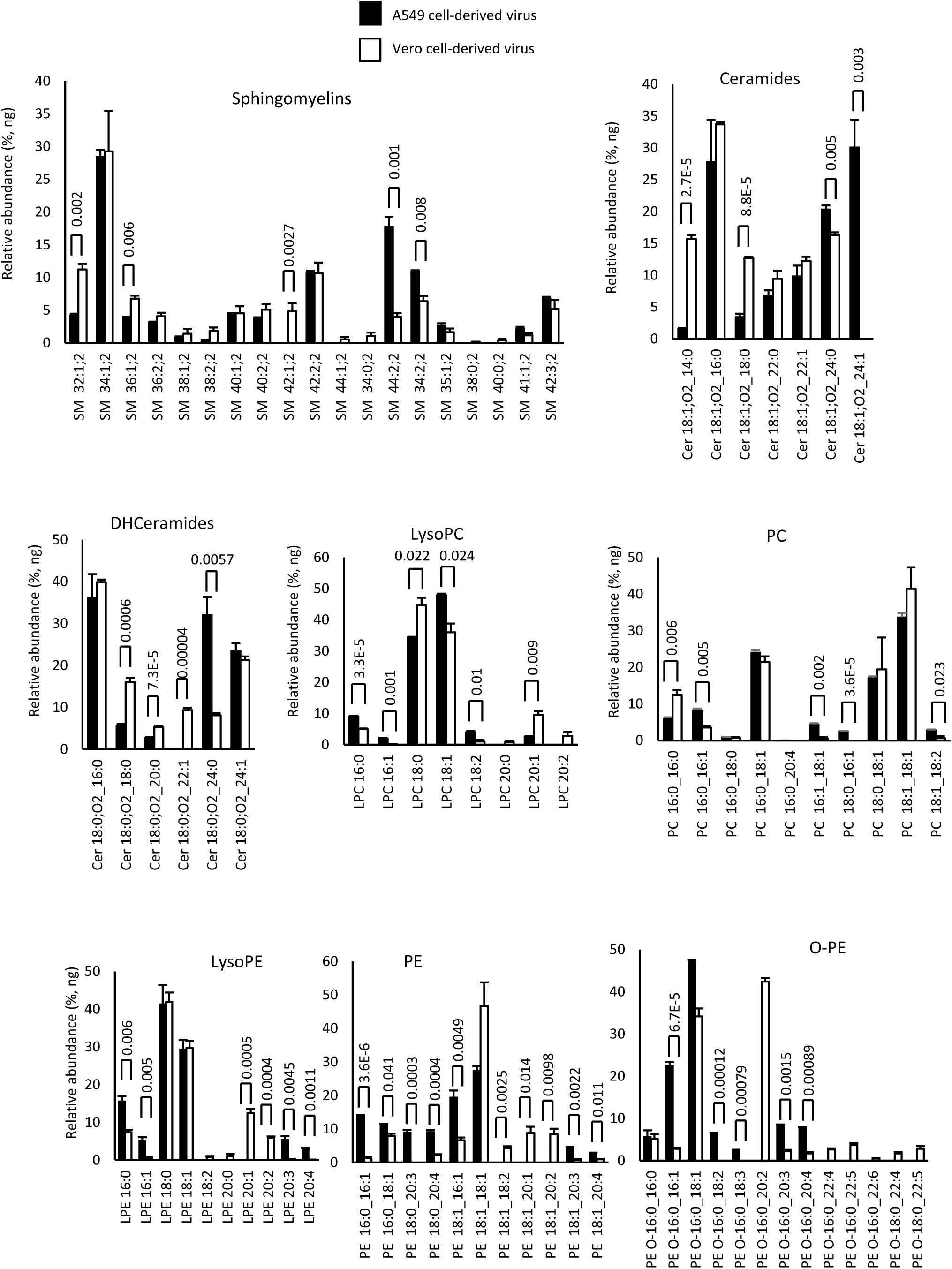
Comparison of lipid molecular species detected in SARS-CoV2 derived from Vero or A549 cells shows some cell-dependence in FA composition across the cell types. Lipids were totalled within each category (ng) then expressed as % for n = 3 preparations/analyses, mean +/- SEM, as outlined in Methods. Unpaired Students T test, followed by Benjamini Hochberg correction where there were >20 variables.

**Figure 3.**
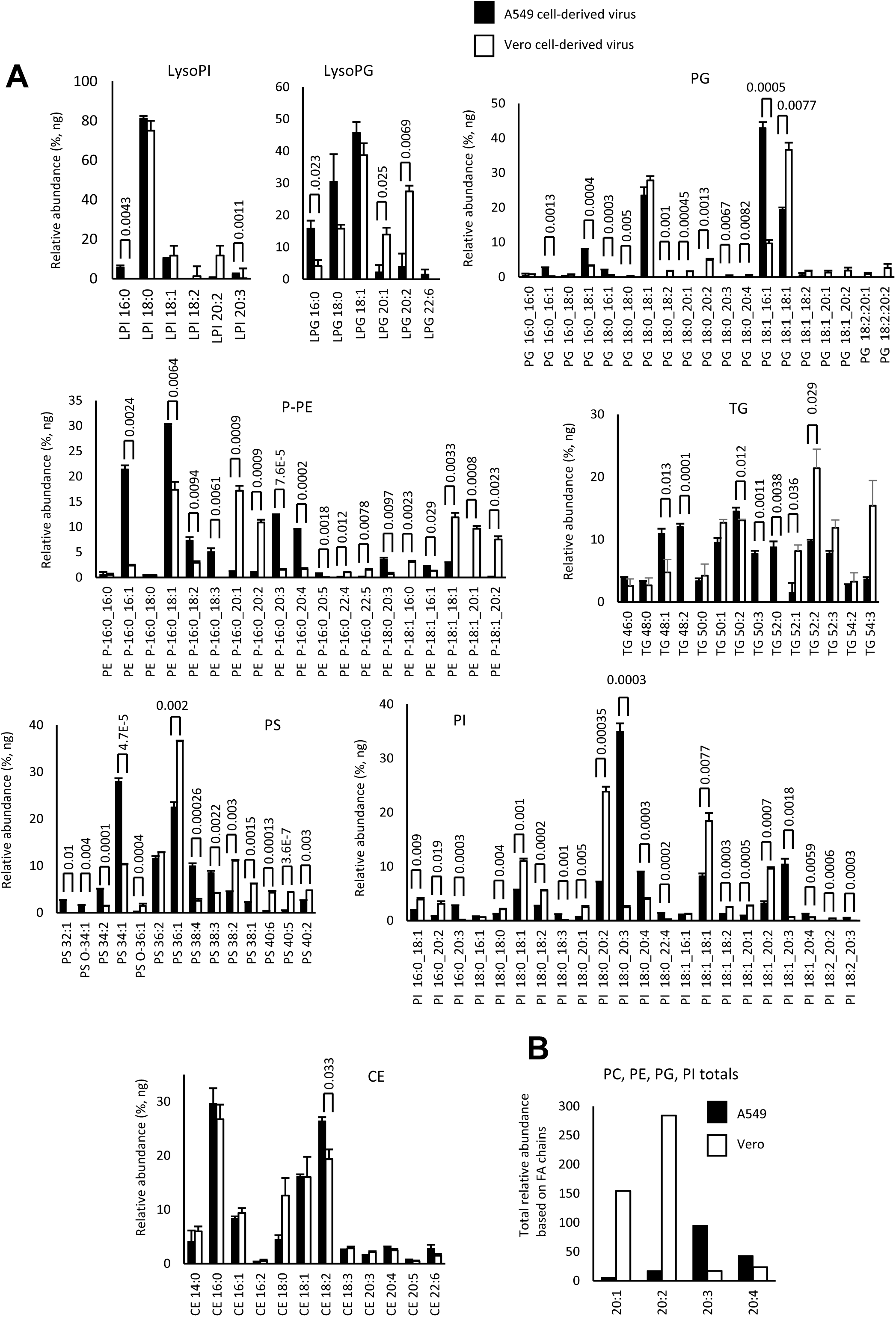
Comparison of lipid molecular species detected in SARS-CoV2 derived from Vero or A549 cells shows some cell-dependence in FA composition across the cell types. *Panel A*. Lipids were totalled within each category (ng) then expressed as % for n = 3 preparations/analyses, mean +/- SEM, as outlined in Methods. Unpaired Students T test, followed by Benjamini Hochberg correction where there were >20 variables. *Panel B*. Species of PL (PC, PE, PI, PG) that were determined to contain FA with 20:1, 20:2, 20:3 or 20:4 were totalled to generate a comparison for A549 versus Vero cells.

### Generation of an untargeted lipidomics dataset for future lipid mining

Next, to generate an untargeted dataset, virus lipids from Vero cultures were separated using HILIC chromatography in triplicate, subtracting extracted blanks, scanning from 50-2000 amu, at 20K resolution. Retention time windows were identified using internal standards for PC, PE, PG, PI, lysoPC, lysoPE, lysoPG, lysoPI, glycerides, SM and Cer. Since electrospray ionisation high resolution mass spectroscopy (ESI-HRMS) runs contain large numbers of artefactual ions including in source fragments, isotope peaks, common contaminant ions in blanks, salt clusters, ion stacks and other features, two informatics approaches were applied to clean up the dataset. First, XCMS was used to align and integrate peaks, before a full clean up using LipidFinder 2.0 was performed(*36, 37*). LipidFinder is designed to remove as many of these artefacts as possible while retaining real lipids, and also subtracts blank signals to correct for background. Large numbers of ions were returned in the dataset and putative matches are provided from the LIPID MAPS Structure Database (LMSD) Bulk search, mapped to the LIPID MAPS classification (*38, 39*) (Supplementary Data2.xls). Using internal standards, we isolated ions matched to specific categories as described above, and putative matches that fell outside retention time windows were moved to the Unknown category, and names removed. Since this analysis is based on precursor mass only, bulk annotation should be used, and putative matches are provided as unvalidated examples of potential structure, where shorthand annotation is not available (e.g. prenols, polyketides, saccharolipids)(*39*). We recommend if this data is to be used that investigators rigorously confirm structures of interest using MS/MS and we would be able to provide lipid extracts on request in order to help such an endeavour.

Overall, our lipidomics data establish SARS-CoV2 membrane as highly enriched in PL, particularly PC, PE and PI, but with rather low levels of cholesterol, PS and SM. They also show that the membrane is influenced to some extent by the host cell of origin in relation to specific molecular species of lipids detected.

### The SARS-CoV2 lipid membrane external leaflet is unable to maintain asymmetry of phosphatidylethanolamine (PE) and phosphatidylserine (PS)

Mammalian membranes maintain asymmetry via the action of flippases and floppases which retain aPL on the inner membrane (*40*). Here, we determined the proportion of PE and PS molecular species on the surface of viral particles using derivatisation-LC/MS/MS (*41*). Adding together the molecular species measured, the external levels of aPL were 48% or 52% for virus from A549 or Vero cells respectively. However, for Vero cells, the % of PS externalised was consistently lower, around 27%, versus 56% for A549, with the level of PS 18:1_18:1 being significantly reduced (Figure 4 A). For PE, the overall external levels were 52% for both cell types. Generally, the pattern of external aPL was similar for both virus preparations, with around half of the aPL being exposed on the external leaflet (Figure 4A). Thus, unlike mammalian cells, SARS-CoV2 particles are unable to maintain asymmetry of aPL. As a comparison, we previously showed that when platelets are thrombin activated, calcium-dependent scramblase externalises PE/PS, to only around 3-4 mol%, from around 0.2-0.5% basally(*40*). This is sufficient to support complete binding and activation of coagulation factors, leading to haemostasis and thrombosis(*40*). Thus, considering the total virus levels of PE and PS (Figure 1 C), with the proportions detected externally (Figure 4 A), these particles will expose some external facing PS, along with very high levels of external PE, far higher than would be present on platelets during physiological or pathological haemostasis.

**Figure 4.**
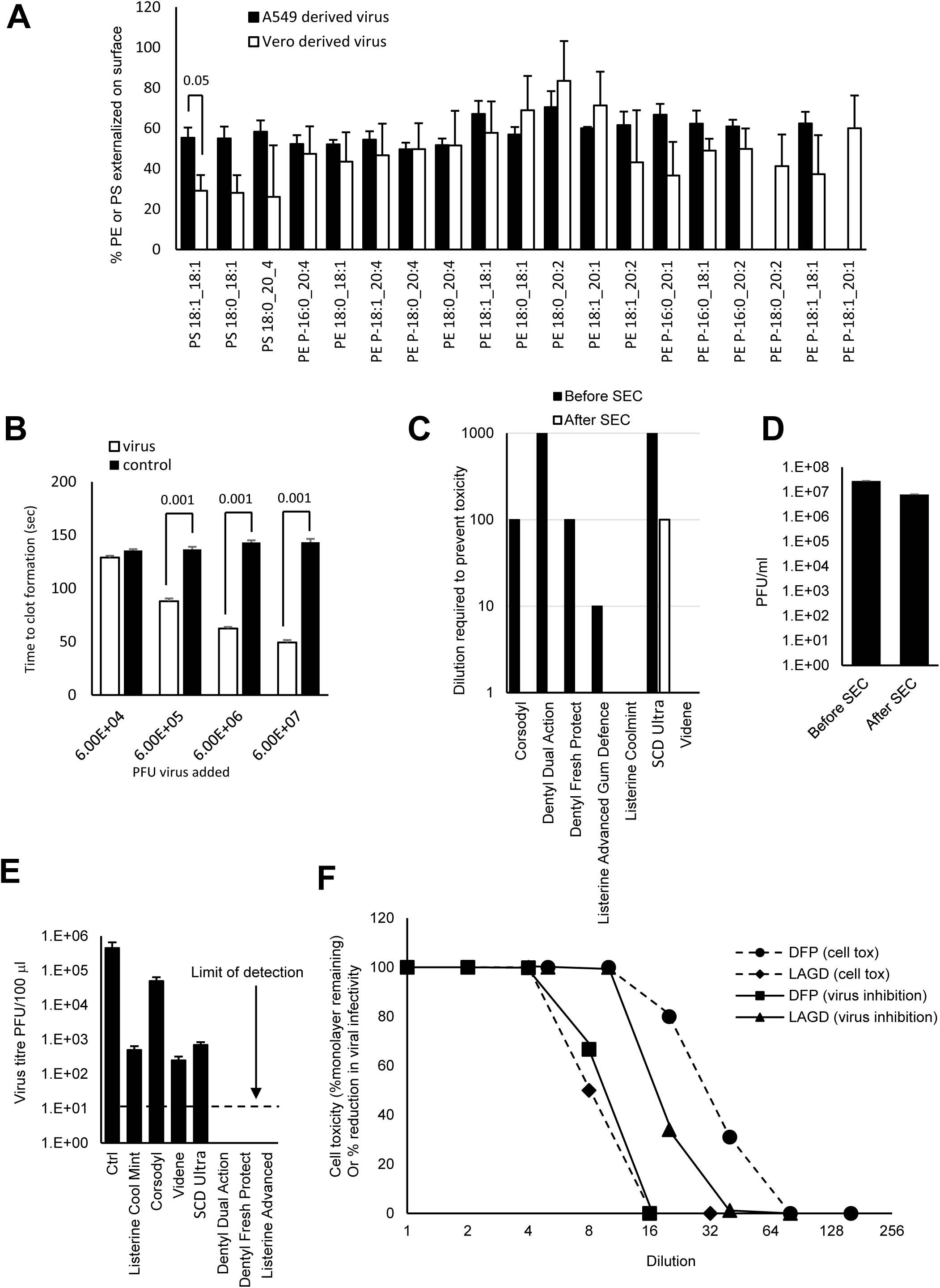
SARS-CoV2 membranes externalise large proportions of PE and PS on the surface of the particles, virus can enhance plasma coagulation, and virions are sensitive to inactivation by surfactants in widely available oral rinses beyond the level required for EN14476 standard. *Panel A. High external exposure of aPL on the surface of SARS-CoV2*. External PS and PE were determined as described in Methods for 3 preparations of SARS-CoV2 (n = 3, mean +/- SEM) using virus from A549 or Vero cells as indicated, unpaired Students T test. *Panel B. Virions enhance plasma coagulation*. Virus was added to normal human plasma as outlined in Methods, and time to gel/clot formation measured. PBS was added to control samples. n=3, +/- SEM, one way ANOVA with Tukey Post Hoc test. *Panel C. Size exclusion chromatography (SEC) can remove mouthwash to prevent any direct impact on cell viability during infectivity testing*. Mouthwashes were mixed with DMEM and synthetic salivary secretions, then 100μl of the mixture purified through a S-400 HR spin column, diluted by serial 10-fold dilution in DMEM/10, and inoculated onto VeroE6/ACE2/TMPRSS2. After 72h, overlays were removed and monolayers were fixed and stained with crystal violet, then toxicity was scored based on visual inspection of monolayer integrity (mean, n = 2, representative of 3 independent experiments). *Panel D. Removal of mouthwash using SEC has little impact on viral infectivity*. 100μl virus was purified through a S-400 HR spin column, and live virus measured by plaque assay on VeroE6/ACE2/TMPRSS2 (n = 3 – 4, mean +/- SEM). *Panel E. Several mouthwashes can significantly reduce infectivity, while some totally eradicate the virus, achieving the EN14476 standard*. Virus was mixed with synthetic salivary secretions and mouthwash, then purified by SEC after 30 seconds, before being titrated by plaque assay on VeroE6/ACE2/TMPRSS2 as described in methods (n = 2, mean +/- SD, representative of 3 independent experiments). *Panel F. Comparing selectivity for virus inactivation versus host cell toxicity reveals differential effects*. For cell toxicity, serial 2-fold dilutions of dental fresh protect (DFP) or Listerine Advanced Gum Defence (LAGD) were made, then added to VeroE6/ACE2/TMPRSS2 monolayers for 30 seconds, washed off, and replaced with media. 3 days later, monolayers were stained with crystal violet and scored for toxicity. For virus infectivity, serial 2-fold dilutions of mouthwashes were made, then incubated with SARS-CoV2 and a soil load for 30 seconds. After purification by SEC, samples were titrated by plaque assay on VeroE6/ACE2/TMPRSS2. Inhibition was calculated relative to virus incubated with media alone (n = 1 (virus toxicity) or 2 (cell toxicity, mean), representative of 3 independent experiments).

### SARS-CoV2 virions enhance plasma coagulation

Given the key role of external facing PE and PS in together supporting blood clotting, we next tested whether virus could regulate the ability of plasma to coagulate *in vitro*. Gradient purified virions were added to plasma in the presence of CaCl_2_ and the Activated Partial Thromboplastin Time (APTT) was measured as outlined in Methods. Here, the presence of a glass surface stimulates the “contact” or intrinsic pathway, resulting in a cascade of factor activation and eventually fibrin clot formation. Virions dramatically reduced the time taken for clot formation in a concentration dependent manner, at minimum concentrations of ∼6×10^5^ PFU/ml (Figure 4 B). Although data is not available on blood virus levels in severe disease, we note that the levels able to enhance clot formation are well within the range of levels detected in saliva, BAL and subglottic aspirates in patients, who frequently carry loads of 10^6^-10^7^, and can even be above 10^8^ PFU/ml (*42*).

### Lipid-disrupting oral rinses reduce viral infectivity in vitro achieving EN14476 virucidal standards

Having characterised the envelope composition, we next investigated whether it was possible to disrupt the lipid envelope using oral rinses that have been designed to be antimicrobial, but also contained constituents potentially capable of targeting generic PL based membranes. To define biological activity, we assessed viral infectivity *in vitro*, in the presence of a soil load to mimic the components of the nasal/oral cavity. To examine the activity of a range of products, seven formulations were tested (Table 2) including rinses containing cetylpyridinium chloride (CPC, Dentyl Dual action, Dentyl Fresh Protect, SCD Ultra), chlorhexidine (Corsodyl), ethanol/ethyl lauroyl arginate (LAE) (Listerine^®^Advanced Defence Gum Therapy), ethanol/essential oils (Listerine^®^Cool Mint) and povidone iodine (Videne). The impact of a 30 second exposure of virus to rinse formulation was assessed by plaque assay.

**Table 2.** Formulations of mouthwash products used in the study.

| Product | Active Ingredients | Other active ingredients |
| --- | --- | --- |
| **Corsodyl** | 7 % (v/v) ethanol, 0.2 % (w/v) chlorhexidine | Peppermint oil |
| **Dentyl Dual Action** | 0.05 %-0.1 % (w/v) cetylpyridinium chloride | isopropyl myristate, Mentha Arvensis extract |
| **Dentyl Fresh Protect** | 0.05 %-0.1 % (w/v) cetylpyridinium chloride | xylitol |
| **Listerine® Cool Mint** | 21 % (v/v) ethanol | thymol 0.064 %, eucalyptol 0.092 %, methyl salicylate 0.060 % and menthol 0.042 % (all w/v) |
| **Listerine® Advanced Defence Gum Treatment** | 23 % (v/v) ethanol | ethyl lauroyl arginate HCl (LAE) 0.147% w/v |
| **SCD Ultra** | 0.07-0.1 % (w/v) cetylpyridinium chloride (0.05-0.1%), sodium citrate - citric acid 0.15 %, sodium benzoate 0.1 % (all w/v) | sodium monofluorophosphate |
| **Videne** | 7.5% iodinated povidone equivalent to 8.25 mg/ml iodine |  |

The assay was optimised to: (i) exclude potential for mouthwash to interfere with plaque assay through direct toxicity towards host cells, (ii) prevent persistence of effect on virus beyond the 30 sec exposure time and (iii) consider the choice of soil load to best model human oropharynx conditions. An important refinement was the use of VeroE6 which stably overexpress ACE2 and TMPRSS2. This significantly improves viral infectivity, with SARS-COV2 entering >1log more efficiently than parental VeroE6, significantly enhancing assay sensitivity (*35*). Rather than BSA alone, our soil load comprised mucin (type I-S), BSA and yeast extract (as in (*29*)) to better mimic the charged polymeric mucin matrix lining the oral and nasal mucosa. Mucin type I-S is generated in salivary glands and interacts with oral mucosa, food and microbiome. To exclude a direct impact of mouthwash on cells, host cell viability was measured with/without the addition of dilutions of mouthwash for 1 hr (the time taken to infect the cells with SARS-CoV2), in the absence of virus, but the presence of soil load. Five of the seven products reduced cell viability when added undiluted. This cytotoxicity was concentration dependent and reduced via serial-dilution (Figure 4 C). To address this problem, size-exclusion chromatography (SEC) was employed to remove mouthwash from virus prior to plating on cells. This also ensured that anti-viral activity did not continue while virus was diluted and titrated. Purification of the virus on S-400 HR Microspin^®^ columns under control conditions (no mouthwash), resulted in minimal (3.5-fold) loss of infectivity (Figure 4 D). When mouthwashes (without virus) underwent SEC, the flow-through was non-toxic against the cell monolayer for all products with the exception of SCD Ultra (Figure 4 C). SEC was therefore used for all in vitro mouthwash testing. These optimisations enabled the detection of a >5-log10 decrease in virus titre, with the exception of SCD Ultra for which a >4-log10 decrease was measurable. This is above the 4-log10 reduction in activity specified by EN14476, allowing the testing of mouthwash to international virucidal standards, as detailed below.

Next, the ability of mouthwash to reduce virus infectivity, after a 30-second exposure in a soil load, was tested using the optimised plaque assay. Two CPC-containing mouthwashes (Dentyl Dual Action, Dentyl Fresh Protect) and a mouthwash containing 23% v/v ethanol/LAE (Listerine^®^ Advanced DGT) eradicated the virus completely, giving >5-log10 reduction in viral titres, and thus met EN14476 as a virucide. In contrast, only moderate effects (∼3-log fold reduction) were observed with PVP-I (Videne), CPC/sodium citric acid/benzoate (SCD Ultra) and 21 % v/v alcohol/essential oils (Listerine^®^ Cool Mint) (Figure 4 E), which failed to meet EN14476. Chlorhexidine (Corsodyl; <2 log fold reduction) was least effective.

### Oral rinse formulations exhibit differential selectivity in virus- and host cell inactivation

For products with antiviral activity, it is relevant to determine selectivity for the virus as opposed to host cells, since potential toxicity *in vivo* should be considered. We showed that the SARS-CoV2 membrane is similar to ER/Golgi in terms of PL composition (Table 1), and unlike plasma membrane is extremely low in cholesterol and SM. However, whether this is sufficient to reveal differential impacts of oral rinses needed to be experimentally determined. We compared the sensitivity of VeroE6 cells with SARS-CoV2 virions, to dilutions of the two formulations showing the highest efficacy – i.e. CPC (Dentyl Fresh Protect) or ethanol/LAE (Listerine^®^Advanced DGT) following 30 second exposure in the presence of soil load (Figure 4 F). In vitro cell toxicity varied 8-fold between the virucidal mouthwashes (Dentyl Fresh Protect and Listerine^®^ Advanced DGT). Listerine^®^ Advanced DGT showed higher selectivity for virus over cultured cells than Dentyl Fresh Protect, as shown by calculated IC50s (Figure 5 A) which was approximately 2 times more potent at inactivating virus (Figure 4 F). Thus, while neither product demonstrated a high selective index for virus versus cells, the SARS-CoV2 envelope lipid composition may, in principle, enable selection of more targeted formulations with lower impact on host cells.

**Figure 5.**
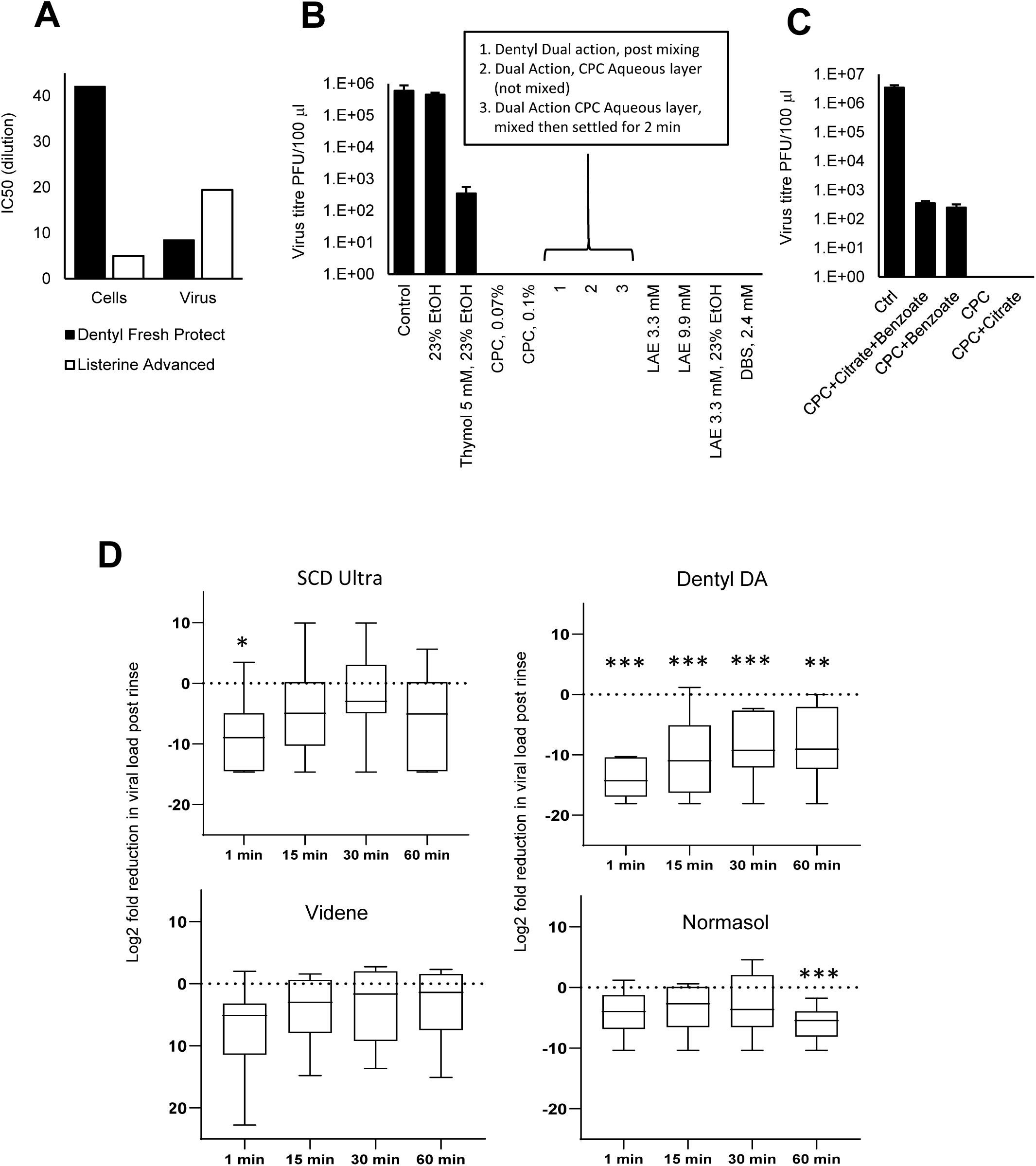
IC50 values for cell versus virus toxicity, antiviral efficacy of mouthwash components, and SARS-CoV2 salivary load is differentially reduced following a brief oral rinse. *Panel A. IC50 values were calculated from data in Figure 4, Panel F using Graphpad Prism. Panels B,C. Surfactants are responsible for the highest virucidal activity in mouthwash formulations*. 100μl virus was mixed with synthetic salivary secretions and the indicated components from different mouthwash formulations, for 30 seconds. Virus was purified through a S-400 HR spin column, and live virus measured by plaque assay on VeroE6/ACE2/TMPRSS2. (n = 2, representative of 3 independent experiments). *Panel D. Oral rinsing significantly impacts salivary viral load*. Samples of saliva were obtained prior to a 30 sec rinse with a mouthwash, and then at various time intervals post rinse, as described in Methods. Saliva was tested for the presence of infective virus using an infectivity assay as described (n = 7,8,6,6 for SCD Ultra, Dentyl Dual Action, Videne and Normasol, respectively). Log2fold reduction in PFU/ml saliva post rinse is shown at the various time points tested. Data is shown as box and whisker plots with median, intraquartiles and range shown. *** p<0.005, **p<0.01, unpaired t-test, for each time point compared with pre-rinse values.

### Surfactants in oral rinses provide the strongest antiviral effects

Despite all mouthwashes containing antibacterial compounds, they demonstrated widely varying abilities to inactivate SARS-CoV2, indicating that careful selection for clinical use may be important. To determine which components were responsible for this activity, SARS-CoV2 was exposed to active constituents (alone or combined) from the relevant rinses, using concentrations found in individual formulations (Figure 5 B, Table 2). CPC, the active component in Dentyl Fresh Protect, eradicated live virus at both concentrations tested (Figure 5 B). Dentyl Dual Action contains CPC/isopropyl myristate (IPM) in a biphasic aqueous-oil system that requires shaking before use. CPC is predominantly in the aqueous phase, while IPM is predominantly in the oil layer. The fully-shaken rinse completely eradicated live virus (Figure 5 B). The aqueous CPC layer (without prior mixing) was also effective, as was the aqueous layer obtained after shaking followed by 2 min settling (to ensure IPM saturation of the CPC layer) (Figure 5 B). Thus, CPC alone can eradicate SARS-CoV2 and IPM isn’t required.

Listerine^®^Advanced DGT contains ethanol at 23 % v/v and LAE (3.3 mM), while other formulations (e.g. Listerine^®^Cool Mint) contain ethanol with essential oils: thymol, menthol and eucalyptol. Whilst 23 % v/v ethanol alone had no consistent impact, the addition of thymol (5 mM) resulted in a 3-log reduction in virus titres (Figure 5 B). This indicates that Listerine^®^Cool Mint reduces virus titres due to the essential oils, with ethanol mainly providing oil solubility. Aqueous solutions of LAE below (3.3 mM) and above (9.9 mM) the critical micelle concentration (cmc, 4.9 mM (*43*)), completely eradicated SARS-CoV2, mirroring the potent anti-viral activity of Listerine^®^ Advanced DGT, which contains 3.3 mM LAE (Figure 5 B). This was seen with or without 23 % ethanol inclusion, indicating that LAE is responsible for the antiviral activity of this product. To determine the potential effect of charge on molecular interactions with the viral lipid membrane, in addition to CPC and LAE (cationic surfactants), the effect of the anionic surfactant dodecylbenzensulfonate (DBS) was tested and found to completely eradicate infectivity (Figure 5 B).

One mouthwash (SCD Ultra) showed only a 3-log reduction in virus titres despite containing CPC (Figure 4 E). This formulation also contains citrate and benzoate. When these were separately added to CPC, citrate had no effect, however benzoate reduced the ability of CPC to kill virus (Figure 5 C). Therefore, while surfactants such as CPC are essential for antiviral activity, additional components may dampen effectiveness in inactivating SARS-CoV2.

### CPC-containing mouthwashes reduce the salivary viral load of SARS-CoV2 in COVID 19 patients

Although a subset of mouthwashes were effective *in vitro*, it was important to determine their effectiveness *in vivo*, where virus is being shed continually in the oropharynx. A randomised clinical trial was undertaken to measure the antiviral efficacy of mouthwashes following a 30 second rinse. 78 hospital in-patients with PCR-diagnosed COVID 19 were recruited, following invitation of over 400 to participate. Despite a positive PCR test in the preceding 14 days, only 27/78 patients had live SARS-CoV2 present in their baseline saliva. Recent studies show that live virus is almost never detected beyond 9 days post-symptom onset in immunocompetent patients (*44*). As our patients were ill enough to be admitted to hospital, many were likely beyond this timepoint. Unfortunately, this was not known at the time sample collection was initiated and only became evident towards the end, with the study terminated at 6 months. By then, new UK daily cases had decreased from 55,892 (31^st^ Dec 2020) to 4,052 (31^st^ March 2021), hospitalised patient numbers were declining and co-morbidity and ventilatory support in these patients rendered them ineligible for randomisation (https://coronavirus.data.gov.uk), making further recruitment impossible. Amongst patients with live virus, saliva was collected before rinsing (baseline), and at 1-, 15-, 30-, and 60-minutes post-rinsing, with mouthwashes containing either containing CPC/IPM (Dentyl Dual Action, n = 8), CPC/benzoate (SCD Ultra, n = 7), PVP-I (Videne, n = 6) or 0.9% w/v NaCl (Normasol, n = 6). Data is shown as both log2-fold reduction from baseline (Figure 5 D) and as individual patient data (Figure 6). Across the entire cohort, baseline salivary viral load varied widely, from 120 PFU/ml to 2.8 × 10^7^ PFU/mL (Supplementary Data1.xls). All four mouthwashes reduced salivary viral load 1-minute post-rinsing, with the smallest reduction being from Normasol^®^ (median 3.9 log_2_ fold reduction from baseline) and the largest Dentyl Dual Action where 6/7 patients recorded no live virus (median 14.3 log_2_ reduction from baseline) (Figure 5 D, Tables 3,4). The persistence of the effects varied with rinse. No significant reduction in salivary viral load was seen with Videne at any of the time-points, while for Normasol^®^ a significant reduction was apparent only at 60 minutes. For SCD Ultra, a significant reduction in viral load was seen at 1 minute only (median 8.9 log_2_ reduction from baseline, Figure 5 D). Dentyl Dual Action was the only product to demonstrate a persistent effect, with a significant reduction evident throughout at 1, 15, 30 and 60 minutes respectively (medians 14.3, 11, 8.8, 9, log_2_ reduction from baseline). Impressively, in 3/8 patients treated with Dentyl Dual Action, no live virus was recovered at any timepoint after the initial rinse (Figure 6).

**Table 3.** Changes in PFU/mL from time 0 to 1, 15, 30 and 60 minutes summarised by geometric mean ratios. Data were analysed using unpaired t test, and

|  | Geometric mean ratio | 95% confidence limits |  | t ratio | p value |
| --- | --- | --- | --- | --- | --- |
|  |  | Lower | upper |  |  |
| Change to 1 minute |  |  |  |  |  |
| Normasol® | 0.0576 | 0.00331 | 1.004 | -2.567 | 0.0502 |
| SCD Ultra | 0.00320 | 0.000059 | 0.174 | -3.520 | 0.013 |
| Dentyl DA | 0.000059 | 0.000008 | 0.000409 | -12.283 | <0.001 |
| Videne | 0.00655 | 0.000016 | 2.648 | -2.153 | 0.084 |
| Change to 15 minutes |  |  |  |  |  |
| Normasol® | 0.0954 | 0.00508 | 1.79 | -2.058 | 0.095 |
| SCD Ultra | 0.0541 | 0.000345 | 8.48 | -1.412 | 0.208 |
| Dentyl DA | 0.000707 | 0.000016 | 0.0317 | -4.510 | 0.003 |
| Videne | 0.0592 | 0.000820 | 4.27 | -1.699 | 0.150 |
| Change to 30 minutes |  |  |  |  |  |
| Normasol® | 0.141 | 0.00318 | 6.187 | -1.334 | 0.240 |
| SCD Ultra | 0.331 | 0.00233 | 47.1 | -0.546 | 0.605 |
| Dentyl DA | 0.00386 | 0.000172 | 0.086 | -4.226 | 0.004 |
| Videne | 0.0970 | 0.00103 | 9.10 | -1.321 | 0.244 |
| Change to 60 minutes |  |  |  |  |  |
| Normasol® | 0.0177 | 0.00219 | 0.143 | -4.964 | 0.004 |
| SCD Ultra | 0.0135 | 0.000108 | 1.69 | -2.181 | 0.072 |
| Dentyl DA | 0.00327 | 0.000098 | 0.110 | -3.854 | 0.006 |
| Videne | 0.110 | 0.000982 | 12.3 | -1.202 | 0.283 |

**Table 4.**
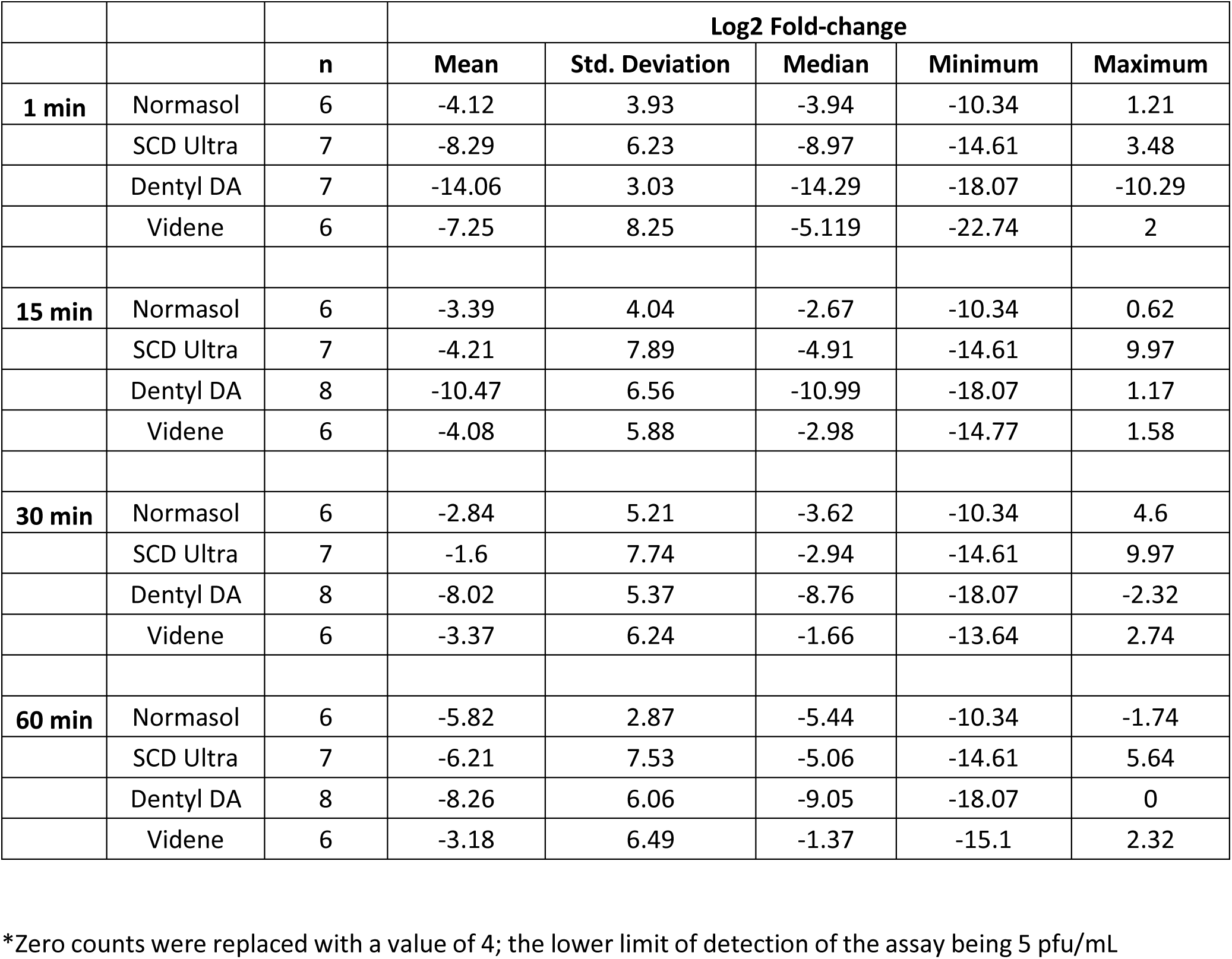
Log2 fold changes of PFU/mL comparing time 0 (baseline) to 1, 15, 30 and 60 minutes, as plotted in Figure 3 A.

|  |  | n | Log2 Fold-change |  |  |  |  |
| --- | --- | --- | --- | --- | --- | --- | --- |
|  |  |  | Mean | Std. Deviation | Median | Minimum | Maximum |
| **1 min** | Normasol | 6 | -4.12 | 3.93 | -3.94 | -10.34 | 1.21 |
|  | SCD Ultra | 7 | -8.29 | 6.23 | -8.97 | -14.61 | 3.48 |
|  | Dentyl DA | 7 | -14.06 | 3.03 | -14.29 | -18.07 | -10.29 |
|  | Videne | 6 | -7.25 | 8.25 | -5.119 | -22.74 | 2 |
| **15 min** | Normasol | 6 | -3.39 | 4.04 | -2.67 | -10.34 | 0.62 |
|  | SCD Ultra | 7 | -4.21 | 7.89 | -4.91 | -14.61 | 9.97 |
|  | Dentyl DA | 8 | -10.47 | 6.56 | -10.99 | -18.07 | 1.17 |
|  | Videne | 6 | -4.08 | 5.88 | -2.98 | -14.77 | 1.58 |
| **30 min** | Normasol | 6 | -2.84 | 5.21 | -3.62 | -10.34 | 4.6 |
|  | SCD Ultra | 7 | -1.6 | 7.74 | -2.94 | -14.61 | 9.97 |
|  | Dentyl DA | 8 | -8.02 | 5.37 | -8.76 | -18.07 | -2.32 |
|  | Videne | 6 | -3.37 | 6.24 | -1.66 | -13.64 | 2.74 |
| **60 min** | Normasol | 6 | -5.82 | 2.87 | -5.44 | -10.34 | -1.74 |
|  | SCD Ultra | 7 | -6.21 | 7.53 | -5.06 | -14.61 | 5.64 |
|  | Dentyl DA | 8 | -8.26 | 6.06 | -9.05 | -18.07 | 0 |
|  | Videne | 6 | -3.18 | 6.49 | -1.37 | -15.1 | 2.32 |

\*Zero counts were replaced with a value of 4; the lower limit of detection of the assay being 5 pfu/mL

**Figure 6.**
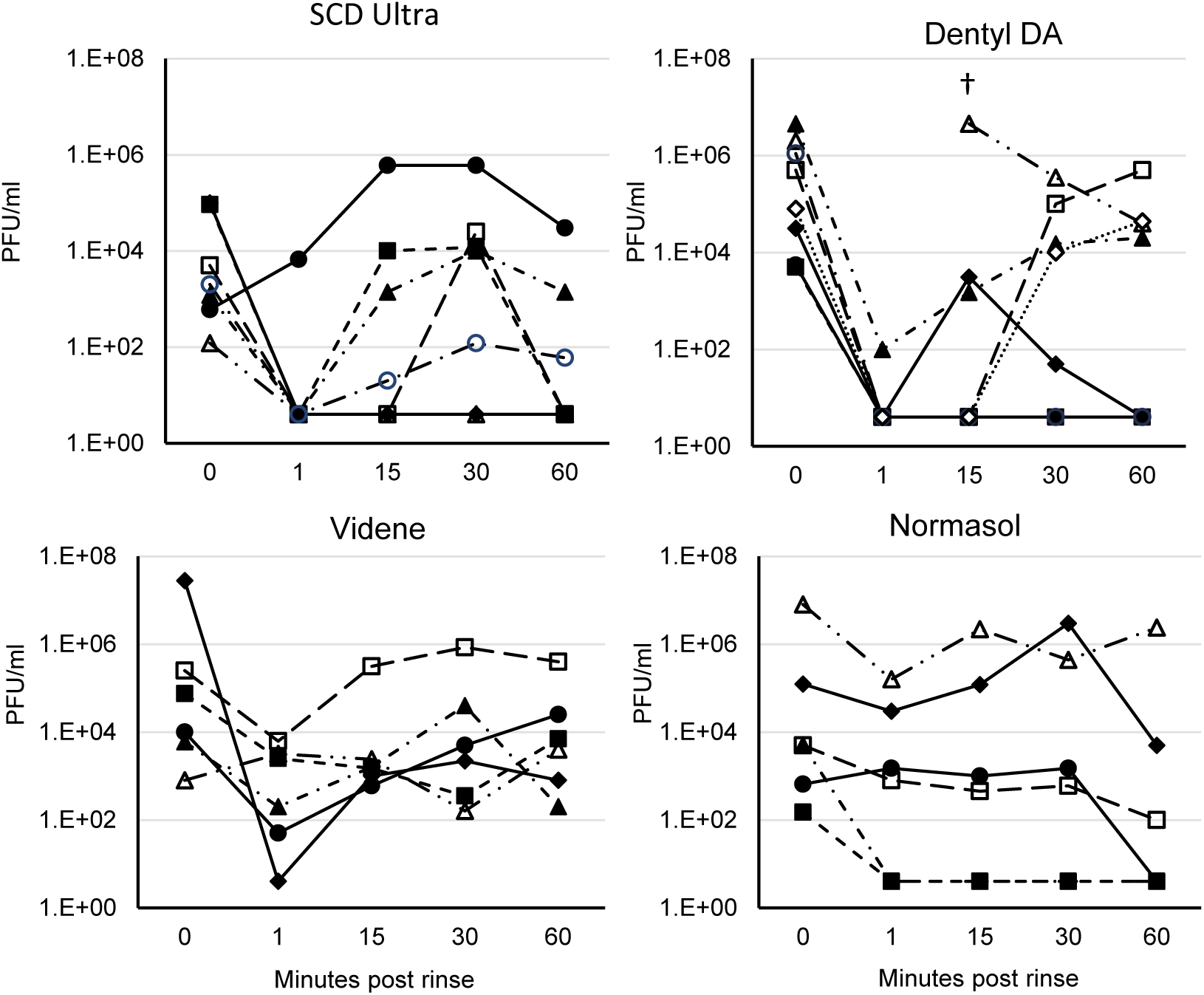
Individual patient data showing how SARS-CoV2 salivary load is differentially reduced following a brief oral rinse. Samples are as outlined in Figure 5. Here, individual data for all participants is shown as PFU/ml plotted as log10. †In this patient administered Dentyl DA, the 1-minute sample had dried before analysis and could not be recorded.

## Discussion

While vaccines and antivirals have targeted the proteins or replication cycle of SARS-CoV2, there has been very little research into the lipid envelope to date. Indeed, there is very little known about lipid membranes of any enveloped viruses, hindering development of strategies targeted directly at the lipids themselves. To address this information gap, a lipidomic analysis of the viral membrane using untargeted and targeted approaches was undertaken. Old studies using thin layer chromatography and total phosphorous analysis, reporting mol% values of rat liver membranes, suggest that PS and SM are enriched in mammalian plasma membranes, versus ER membrane (*9, 45*) (Table 1). On the other hand, PC and PI are enriched in ER vs plasma membrane (*9, 45*). Golgi membranes are generally intermediate between ER and plasma membrane in terms of mol% composition, and are mainly comprised of PC, PE and PI. The cholesterol/PL ratio is highest for plasma membrane, and very low for ER (Table 1). Coronaviruses have long been known to be generated on the ERGIC membrane (*3-8*), however, how this related to their lipid composition was so far unknown.

Here, we found that the SARS-CoV2 membrane is primarily comprised of PC, PE and PI, while having low levels of free cholesterol, PS and SM. This appears most similar to ER, although with even lower proportion of PS, SM and cholesterol than previously reported for that membrane compartment, using older methods. In the mammalian plasma membrane, cholesterol is often concentrated in specialised regions which support receptor dependent signalling, called lipid rafts(*46*). Our data suggest these will be absent from the viral envelope. Furthermore, the cholesterol in plasma membranes regulates fluidity and reduces permeability to small molecules, while SM is also important for reducing fluidity(*47-49*). Thus, the viral envelope and plasma membrane will be very different biophysically. Similarly, bacterial membranes are also considered to be devoid of cholesterol. In this context, this is exploited therapeutically since cholesterol protects host cells from disruption by anti-microbial peptides which directly insert in the bacterial membrane (*50*). Having described the virion envelope in detail, it is now possible to test targeted strategies using liposomes that mimic the SARS-CoV2 membrane, for example by generating liposomes with the exact lipid molecular composition.

Furthermore, we demonstrate that the virion membrane contains lysoPL, from PG, PI, PE and PC, noting that these are known bioactive lipid signalling mediators, and their presence in the envelope could impact on host inflammatory responses to infection. It was recently reported that coronaviruses exit via lysosomal secretion instead of the biosynthetic secretory pathway(*51*). Lysosomes contain high levels of SM and cholesterol (Tabel 1), thus the lack of these lipids indicates that lysosomal passage of virus doesn’t appear to impact envelope composition. Examining the different lipid categories at the molecular species level, a high proportion of saturated/monounsaturated FAs were noted, with little PUFA evident. This most likely reflects the typical FA composition of cultured cells which tend to be lower in PUFA than primary tissues. In human disease, the SARS-CoV2 virus will be actively replicating in oral, nasal and airway epithelia. Studies on airway and tracheal cells have shown their FA composition to be similar to what was seen here, but with significantly more 18:2 and 20:4, which becomes lowered during cell culture (*52, 53*). Thus, virus generated in human airways *in vivo* may have more PUFA than found herein, but this remains to be determined. Notably, we found significant differences in SARS-CoV2 virus lipids, depending on the host cell in which they were generated. This may relate to subtle differences in host cell ER membranes between the cell types. Importantly, inflammation has a significant impact on host cell lipid metabolism, and how this influences virion envelope composition now needs to be tested.

Here, we showed that SARS-CoV2 exposes around 50% of its total molecular species of aPL on the surface of the particle (Figure 4B). As a caveat of the method, membrane proteins or sugars could in theory hinder derivatisation of aPL, and so this value may be a lower-level estimate. In primary cells, energy-dependent processes maintain asymmetry of plasma membranes. This ensures that very low mol% of PE and PS are exposed on the surface, for example, only 3 - 4 % is present on platelets following thrombin activation (*40*). This is because PE and PS promote coagulation, complement binding and uptake of apoptotic cells through their electronegative interactions with Ca^2+^ ions and various proteins (*3, 20, 40*). Although the overall amounts of PS appear to be rather low in virions, PE levels are similar to plasma membrane (Table 1). Thus, exposure of 50 % of aPL on the surface will result in levels of external PE that are around 12-fold-higher than activated platelets(*40*). Indeed, in line with this, we found that purified virions significantly accelerate plasma coagulation *in vitro* (Figure 4 B). In addition, a recent study showed (using a less specific ELISA) that levels of PS on the surface of SARS-CoV2 are sufficient to support PS-receptor dependent viral entry (*17*). Our work extends this significantly by reporting on the ng amounts of PS and PE present, the proportions of PE and PS that are externalised, and the specific molecular species of PE and PS in the membrane. In addition to SARS-CoV2, PS has been implicated in the cellular uptake of several other viruses, thus knowing how much and which molecular species are external facing on the envelope is relevant to other infectious diseases (*10-16*). In summary, our study and others suggest that targeting aPL could support anti-thrombotic, anti-inflammatory or anti-viral strategies for COVID19.

These findings could also be relevant for other enveloped respiratory viruses such as influenza which has long been considered to trigger thrombotic complications of atherosclerosis, including myocardial infarction. Winter peaks in influenza are often followed two weeks later by a peak in ischemic heart disease, hypertension and cerebrovascular disease deaths. Furthermore, many acute vascular events follow upper respiratory infections (reviewed in detail in (*54*)). A recent study found that emergency department visits for respiratory illness were both associated with, and predictive of, cardiovascular disease mortality in adults >65 yrs (*55*). Furthermore, in a meta-analysis, influenza vaccination was associated with lower risk of adverse cardiovascular events (*56*). The mechanisms are unknown and a vascular inflammatory component is likely to play a role. Thus, other enveloped viruses are also strongly associated with thrombotic events, however whether virions themselves directly contribute to coagulation has never been evaluated.

Having determined the composition of the lipid membrane, we next tested the impact of common mouthwash formulations, focusing on surfactants which we reasoned would effectively target a PL-rich membrane. Our data significantly extends other recent studies on enveloped viruses. For example, dequalinium/benzalkonium chloride, PVP-I and ethanol/essential oils reduced SARS-CoV2 infectivity *in vitro* by up to 3-log10 (*29*), while infectivity of HCoV229E was reduced by 3-4-log10 using CPC, ethanol/essential oils and PVP-I (*27, 30*). Also, a moderate (3-log10) antiviral effect of thymol/ethanol is consistent with an *in vivo* study on Listerine^®^Cool Mint against HSV (*57, 58*). However up to now, only one of the products (Listerine^®^Antiseptic, 26.9% ethanol/essential oils) has achieved 4-log10 kill required to pass EN14476 as a virucidal, although this was tested against HCoV229E rather than SARS-CoV2(*30*). Here, we employed live SARS-CoV2 England2 strain, and demonstrated that several mouthwashes (Listerine^®^Advanced DGT, Dentyl Fresh Protect, Dentyl Dual Action) pass EN14476 against this virus.

Importantly, efficacy was not dependent on ‘classical’ antibacterial components of mouthwashes, but instead was critically dependent on the presence of surfactant (CPC/LAE). Whether any of the lipid disrupting components would function in this way was not predictable from the outset; efficacy is determined by both molecular makeup and concentration, with some detergents (e.g. Tween20) not inactivating SARS-CoV2 even at 0.5 %, while others (e.g. Triton X-100) lyse it completely at 0.1 % (*59*). Thus, our finding that components such as essential oils do not eliminate infectivity, is equally important as the finding that CPC and LAE in mouthwashes do. CPC-containing mouthwashes were previously reported to reduce infectivity of other enveloped viruses, including HSV (*60*) and influenza (*61*), while LAE has shown antiviral activity towards HSV-1, vaccinia virus and bovine parainfluenzae 3 (*62*). Thus, our results may be generally applicable to all enveloped viruses. The finding that LAE is virucidal both above and below cmc, and with or without ethanol, suggests that the virucidal activity of LAE is independent of micellar self-aggregation, and that the transfer of individual surfactant molecules into the viral envelope destabilises the bilayer. The ability of CPC alone, at two concentrations above its cmc (1 mM), to fully inactivate SARS-CoV2 is likely due to micelle-forming surfactants having a very different “packing parameter” than the lipids in the viral bilayer (*63*). Mixing the surfactant with lipids may increase the local curvature, causing formation of separate micelles, effectively dissolving the bilayer. Finally, the effect of surfactant was not charge-dependent as both cationic and anionic surfactants were virucidal (Figure 5 B, Figure 7).

**Figure 7.**
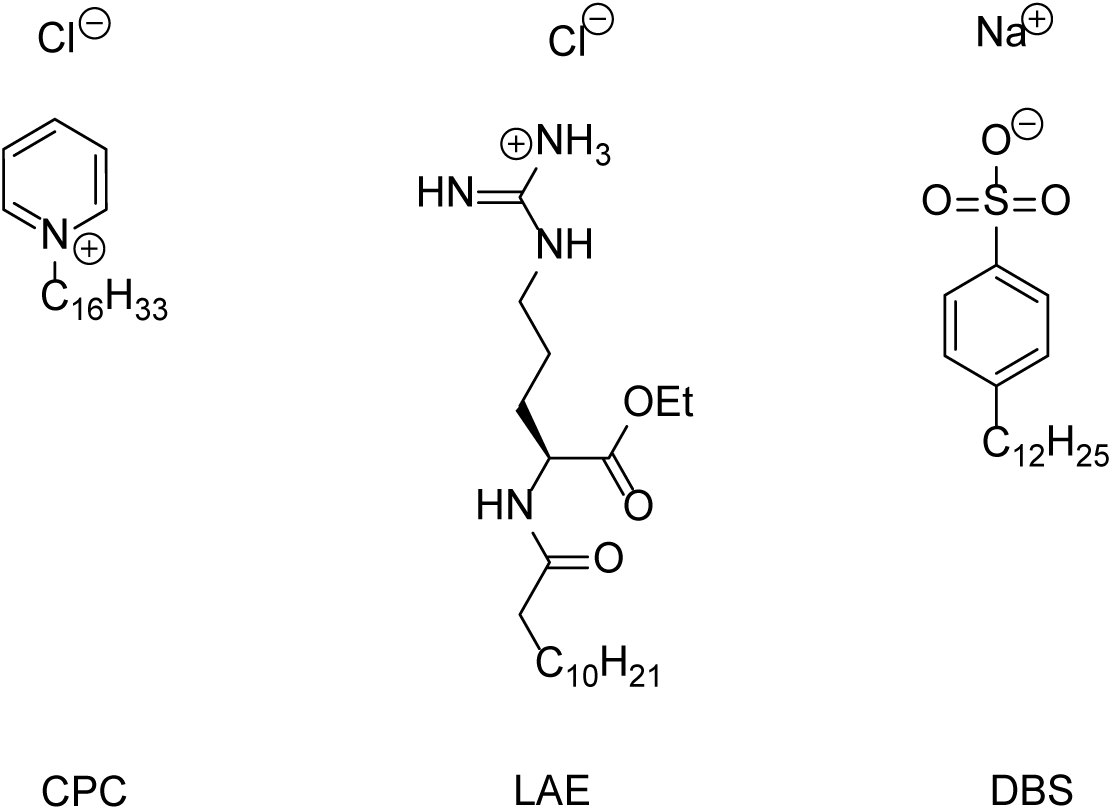
Chemical structure of surfactants used in this study.

Whilst surfactants are highly antiviral in isolation, other mouthwash components may reduce their effectiveness. Benzoate can bind with CPC, dramatically reducing its cmc, without changing its micellar morphology (*64, 65*). Such interactions are typical for combinations of cationic surfactants and aromatic anions (*66*). A reduced cmc indicates a lower concentration of non-aggregated surfactant. The reduction in virucidal effect of CPC caused by benzoate in SCD Max may therefore be due to it reducing the concentration of monomeric CPC. Taken together, our data indicate that whilst product selection is typically made on the basis of individual principal components e.g. CPC, chlorhexidine, or iodine, interactions between ingredients need to be carefully considered.

To address the theoretical potential for toxicity of mouthwash with long term use, we compared cell disruption with viral infectivity and found some minor differences (Figure 4 F). These could be due to the fact that the virus membrane is deficient in some lipids that are known to be enriched in host plasma membrane (e.g. cholesterol, SM). Notably however, >25 % of UK adults use mouthwashes daily with no ill effects reported, while mouthwash was used by almost 200 million Americans in 2020 (*67*) (https://www.statista.com/statistics/286902/usage-mouthwash-dental-rinse-us-trend/). While CPC-containing mouthwashes can show cytotoxicity against monolayers *in vitro*, the lack of observed toxicity *in vivo* likely reflects the complex, differentiated, multicellular nature of primary oral epithelia (*68*). Furthermore, studies have demonstrated the effectiveness of CPC-containing mouthwashes in safely reducing gingival inflammation, despite concerns regarding perturbation of the oral microbiome (*69, 70*). In patients with COVID19, increased disease severity was recently shown to be associated with moderate/severe periodontal disease (*71*). Associations between periodontal inflammation, cytokine release, and altered lipid metabolism have also been established in a range of co-morbidities that associate with poorer COVID19 outcome, including neurodegeneration, diabetes and cardiovascular disease (*72*). Thus, as part of maintaining routine oral health, mouthwash use has potential to impact transmission and disease through both direct and indirect mechanisms.

In many parts of the world, clinical investigations of the oropharynx, including in primary care, dentistry, ENT, and maxillofacial surgery, have been severely curtailed due to the risk of SARS-CoV2 transmission from pre-clinical asymptomatic patients. Here, interventions to reduce the salivary load in patients might be of benefit. Ideally, a large-scale trial would assess the ability of oral rinsing to impact on disease transmission and disease severity however this was not possible in the pandemic situation. Instead, we tested the *in vivo* efficacy of mouthwash on hospitalised ‘moderate’ COVID patients (not requiring intensive medical support or ventilation) and showed a strong impact of surfactant-containing oral rinses on live virus load in saliva. Only three other studies have attempted to address these effects *in vivo* during the pandemic and both were inconclusive due to small numbers of patients, and the use of qPCR rather than live virus titration to determine virus load (*28, 73, 74*).

Our clinical study was designed in the early part of the pandemic, prior to data becoming available showing that throat virus falls to undetectable levels by 9 days post symptom onset (*44*). Thus, our study was challenged by recruitment (inability to provide informed consent and provide salivary samples), co-morbidity and the inability to predict patients with saliva containing live virus (<40% of patients). The study was concluded at 6 months with 78 patients randomised; the largest sample to date of any study, with 27 patients having live virus in their saliva. While all mouthwashes were generally beneficial at 1 minute, the CPC mouthwash (Dentyl Dual Action), was the most effective, significantly reducing live viral load over the entire time course, and completely eliminating it for 1h in multiple patients. An important caveat is that mouthwash use will not target virus in the lower respiratory tract; it remains unclear whether virus that transmits to others arises from the upper or lower respiratory tract, although we have shown that infectious virus is found more commonly in the upper than the lower respiratory tract (*42*). Nevertheless, it remains the case that mouthwashes can destroy enveloped viruses in the oral cavity for sufficient time to enable a dental or oral investigation. Future trials will be required to ensure that this effect is consistent in larger cohorts. However critically, current WHO guidance proposes the use of hydrogen peroxide and PVP-I mouthwashes in dental surgery during the COVID19 pandemic (https://www.who.int/publications/i/item/who-2019-nCoV-oral-health-2020.1). However hydrogen peroxide only weakly inactivates SARS-CoV2 (*75*), and our data shows that surfactant-containing mouthwash is more effective than PVP-I both *in vitro* and *in vivo*. Therefore, there may be a need to re-evaluate this advice.

In summary, we characterise the lipid membrane of SARS-CoV2, as being primarily comprised of PC, PE and PI, with a high proportion of external aPL, and show that the FA molecular species present may vary depending on host cell. Importantly, an inability to maintain asymmetry in the lipid envelope results in live virus being highly pro-coagulant. We also show that surfactant mouthwash that targets the lipid membrane may be a useful component of infection prevention and control strategies for respiratory enveloped viruses (e.g. influenza, SARS, MERS, in addition to SARS-CoV2), during a pandemic. They have the potential to lessen the risk of transmission from asymptomatic carriers to healthcare professionals performing oropharynx investigations, as well as transmission within the wider population, in a similar manner as to how they are being tested against pathogenic oral bacteria(*76-80*). Larger population-based studies are now warranted to determine the impact of this biological effect on transmission. Importantly, the anti-viral actions of oral rinses is not dependent on classical antibacterial components, but instead depends on the sensitivity of the lipid envelope to surfactants as membrane-disrupting agents, which can be made cheaply and easily in LMICS. The membrane of enveloped viruses is likely to remain susceptible to this approach irrespective of mutations that impact vaccine efficacy.

## Materials and Methods

### Aqueous solutions

CPC was dissolved in deionised water at 0.07% or 0.1 (w/v). For aqueous solutions containing LAE, a 33 mM stock solution was prepared by dissolving 693.2 mg of *N*-ethyl lauroyl arginate hydrochloride (Fluorochem, used as received) in deionised water (Elga Purelab Flex), adjusting the pH to 7 using a NaOH solution (pH was determined using a Hanna Instruments pH210 microprocessor pH meter with a VWR simple junction universal combined pH/reference electrode) and making up the solution to 50 mL. The required LAE solutions were then prepared by mixing 1 mL of the stock solution and 9 mL of deionised water (3.3 mM), 3 mL of the stock solution and 7 mL of deionised water (9.9 mM) and 1 mL of the stock solution, 6.7 mL of deionised water and 2.3 mL of ethanol (3.3 mM LAE and 23 vol-% ethanol). For DBS, 807.4 mg 4-dodecylbenzenesulfonic acid, mixture O (Merck Life Sciences UK, used as received) was dissolved in deionised water (Elga Purelab Flex). The pH was adjusted to 6 using a NaOH solution (pH was determined using a Hanna Instruments pH210 microprocessor pH meter with a VWR simple junction universal combined pH/reference electrode) and making up the solution to 1 litre. The solutions of CPC in combination with citrate and benzoate was prepared by dissolving ∼0.11 g of CPC with 0.12 g of citric acid and/or 0.07 g of benzoic acid in deionised water, adjusting the pH as before, and making up to 100 mL. The solution containing deionised water, ethanol and thymol was prepared by dissolving 0.1675 g of thymol in 50 mL of ethanol; 2.3 mL of this solution was combined with 7.7 ml deionised water. All commercial mouthwash preparations are listed in Table 2.

### Cells and viruses

Virucidal assays utilised VeroE6 or A549 cells, a gift from the University of Glasgow/MRC Centre for Virology, UK. To enhance infectivity and produce a more sensitive cell line for detection of virus, both cell types were transduced with lentiviruses encoding ACE2 and TMPRSS2, then drug selected as described (*35*). The England2 strain of SARS-CoV2 was provided by Public Health England, and amplified in VeroE6 cells before being harvested from the supernatant. All cells were grown in DMEM containing 2 % (v/v) FCS, and incubated at 37 °C in 5 % CO_2_.

### Virucidal assays

Virucidal activity of mouthwash was studied in media containing 100 μL mucin type I-S, 25 μL BSA Fraction V, and 35 μL yeast extract to mimic oral secretions. 100 μL of this mixture was added to 100 μL of virus suspension, and 800 μL of the test-product or medium as control. After 30 seconds, virucidal activity was neutralised by 10-fold serial dilution in ice-cold DMEM (containing 10% FCS). Alternatively, virus was purified by size-exclusion chromatography (SEC) to prevent direct cytotoxic effects of the products on the cell monolayer; 100 μL of the mixture was added to a microspin S-400 HR column, and centrifuged for 2min at 700 x *g*. A 10-fold serial dilution was then made of the flow-through in DMEM containing 10% FCS. Virus was titrated by plaque assay; serial dilutions were used to infect VeroE6/ACE2/TMPRSS2 cells for 1 h. Following this, cells were overlaid with DMEM containing 2 % FCS, and 1.2 % Avicel^®^. After 72 h, the overlay was removed, and the monolayer washed and fixed with 100% methanol. Monolayers were stained with a solution of 25% (v/v) methanol and 0.5 % (w/v) Crystal Violet, then washed with water, and plaques were enumerated. For measurement of toxicity, monolayers were similarly incubated, stained with crystal violet, and scored by eye for live cells.

### Harvest of virus particles and lipid extraction for lipidomics profiling

Cells were infected with SARS-CoV2 at MOI=0.01, when cells were 70% confluent, in either serum-free media (Vero cells) or at 2% FCS (A549). At 96h post-infection, supernatants were harvested, cellular debris pelleted (2,000 *xg*, 5min), then virus pelleted through a 30 % sucrose cushion (25,000 rpm, 2.5 h, in a SW28 rotor (112,398 × *g*)). Pellets were resuspended in PBS, purified on a 20 – 60 % sucrose gradient (25,000 rpm, 16 h, in a SW41 rotor (106,882 × *g*)), before being pelleted (35,000 rpm, 1 h, in a SW41 rotor (209,490 × *g*), and resuspended as outlined below. All preparations were analysed for purity and abundance by Nanoparticle tracking analysis using Nanocyte^®^ (Malvern Panalytical), and by Western blot. For PS externalisation, samples were used immediately. For lipidomic profiling they were used immediately or stored for a few days at -80 ° as snap frozen pellets.

For untargeted and targeted lipidomics, virus particles were resuspended in 0.5 ml of PBS, which was then spiked with 10 μl Splash mix (Avanti Polar Lipids), containing : d18:1-18:1(d9) SM (296ng), 15:0-18:1(d7)PC (1.506μg), 15:0-18:1(d7)PE (53ng), 15:0-18:1(d7)PG (267ng), 15:0-18:1(d7)PI (85ng),

18:1(d7) Lyso PC (238ng), 18:1(d7) Lyso PE (49ng), cholesterol-d7 (984ng), CE 18:1-d7 (3.291μg) TG 15:0/18:1-d7/15:0 (528ng), 15:0-18:1(d7) PS (39ng) and 20ng of 17:1 Lyso PG, 17:1 Lyso PI. Samples were also spiked with 5μl of ceramide/sphingoid internal standardmixII (Avanti Polar Lipids) containing 56.99ng of d18:1/12:0 Ceramide. Samples were then extracted using a Bligh and Dyer method. Briefly, 1.9 ml of solvent mixture 2:1 methanol:chloroform v:v was added to 0.5 ml sample. Samples were vortexed for 30 sec, and then 0.625 ml of chloroform added. Samples were vortexed again (for 30 sec) and 0.625 ml of water then added. Samples were vortexed for 30 sec and centrifuged at 1500 rpm, at 4 °C, for 5 min. Lipids were recovered from the lower layer, and evaporated to dryness using a Labconco RapidVap^®^. Extracted lipids were reconstituted in 200 μl methanol and stored at -80 °C until analysis.

### Targeted LC/MS/MS analysis of lipid categories and classes

Targeted assays were performed on 3 separate culture preparations of gradient purified SARS-CoV2 virus, from either Vero or A549 cells *(i) Vero*: For Preps 1 and 2, three technical replicates of the same extracted lipid samples were analysed and averaged to give one set of mean values per Prep (n = 1 per prep). For Prep 3, three separate extractions and analyses were performed (different virus particles in each extraction) and then values averaged to give one value per lipid (n = 1). Combining Preps 1-3 gave n = 3 values for lipid molecular species. Standards for Ceramides (Cer) and DHCeramides (DHCer) were only included in Prep 3, although these lipids were detected as present in all preparations. Data for Cer and DHCer are from 3 separate virus isolate extractions using Prep 3. *(ii) A549*. All samples had Cer and DHCer standards included. Prep 1 was generated from an individual isolate, while Preps 2 and 3 arose from a larger scaled up culture preparation, separated into Preps 2 and 3, prior to lipid extraction. Preps 1-3 were analysed, giving n = 3 which was averaged to generate mean values. A full list of all lipids analysed is shown in Supplementary Table 1. Category specific figures show lipid molecular species comprising at least 2% of the signal of the most abundant lipid in that class. We note that PEs annotated as plasmalogen (vinyl ether) could also include isobaric ether lipids.

HILIC LC-MS/MS was used for PLs and sphingolipids (SL) on a Nexera liquid chromatography system (Shimadzu) coupled to an API 6500 qTrap mass spectrometer (Sciex). Liquid chromatography was performed at 35 °C using a Waters XBridge Amide column, 3.5μm, 4.6 × 150 mm, at a flow rate of 0.7 mL/min over 24 min. Mobile phase A was (water/acetonitrile 5/95; v/v and 1 mM ammonium acetate) and mobile phase B was water/acetonitrile (50/50; v/v and 1 mM ammonium acetate). The following linear gradient for B was applied: 0.1 % B – 6 % B over 6 min, 6 – 25 % B over 4 min, 25 – 98 % B over 1 min, 98 – 100 % B over 2 min. At 13.5 min, the flow rate changed to 1.5 ml/min and remained at 100 % B until 18.7 min where it returned to 0.1 % B. Flow rate then returned to 0.7 ml/min at 23.5 min. Source conditions for positive mode were IS 5.5 kV, CUR 35, TEM 550°C, GS1 50, GS2 60. Negative mode source conditions were IS -4.5kV, CUR 35 psi, TEM 550°C, GS1 50 psi, GS2 60 psi. Dwell time was calculated in Analyst automatically based on the number of MRMs. This is a scheduled method with pos/neg switching throughout. PLs and ceramides were quantified using an external calibration with the following standards, based on a single standard per class (Splash mix) since structurally related lipids tend to closely elute on HILIC chromatography: d18:1-18:1(d9) SM, 15:0-18:1(d7)PC, 15:0-18:1(d7)PE, 15:0-18:1(d7)PG, 15:0-18:1(d7)PI, 18:1(d7) Lyso PC, 18:1(d7) Lyso PE, 17:1 Lyso PG, 17:1 Lyso PI. PC’s, PE’s, PI’s, PG’s Lyso PG’s, Lyso PI’s, Lyso PE’s and Lyso PC’s were quantified from standard curves containing two primary standards each (with the exception of Lyso PE and Lyso PG which had one primary standard each) (PC 16:0-18:1, PC 18;0-22:6, PE 16:0-18:1, PE 18:0-20:4, PG 16:0-18:1, PG 18:0-22:6, PI 16:0-18:1, PI 18:0-20:4, Lyso PC 16:0, Lyso PC 18:0, Lyso PE 16:0, Lyso PG 16:0, Lyso PI 16:0, Lyso PI 18:0). Ceramides were calculated from a standard curve generated by serially diluting the internal standard. Sphingomyelins were calculated based on the following equation: (Area A/Area IS) * (ng IS added). For confirming the absence of serum contamination of lipids in purified virus cultured from A549 cells, blank isolates (medium+ 2% serum) were extracted and then analysed using direct injection precursor scanning MS/MS for the presence of PE (prec 196, -ve ion mode), PC (prec 184, +ve ion mode) and CE (prec 369, +ve ion mode), comparing with virus lipid extracts.

Phosphatidylserine (PS) does not resolve well using HILIC chromatography and was instead analysed using a shotgun method to generate bulk species data. A neutral loss scan (NL 87) was acquired in negative ion mode on the Sciex 6500 platform, to obtain a list of precursor PS species present in the virus lipids. Samples were injected (10 μl for Preps 1,3, 5μl for Prep 2) under flow (mobile phase: methanol + 1 mM Ammonium acetate, 0.2 ml/min), with source and MS conditions as follows: CUR 35, IS -4500, TEM 500, GS1 40, GS2 30, DP -50, CE -36, CXP -29. Once the main PS species were identified, a multiple reaction monitoring (MRM) approach was used, monitoring precursor ions (as determined by the NL precursor scan), to the NL fragment of *m/z* 87. These were quantified against 15:0-18:1(d7) PS, present in the splash mix. MRMs were as follows: *m/z* [M-H]^-^ 758.6-671.5 (PS 34:2), 760.6-673.5 (PS 34:1), 774.6-687.5 (PS O-36:1), 786.6-699.5 (PS 36:2), 788.6-701.6 (PS 36:1), 810.7-723.6 (PS 38:4), 812.7-725.6 (PS 38:3), 814.7-727.6 (PS 38:2), 816.7-729.6 (PS 38:1), 834.7-747.6 (PS 40:6), 836.7-749.6 (PS 40:5), 842.7-755.6 (PS 40:2).

LC-MS/MS for free cholesterol and cholesterol esters (CE) and LC-MS analysis of triacylglycerides (TG) was performed on a Nexera liquid chromatography system (Shimadzu) coupled to an API 4000 qTrap mass spectrometer (Sciex). Liquid chromatography was performed at 40 °C using a Hypersil Gold C18 (Thermo Fisher Scientific) reversed phase column (100 × 2.1 mm, 1.9 μm) at a flow rate of 0.4 mL/min over 11 min. Mobile phase A was (water/solvent B 95/5; v/v and 4 mM ammonium acetate) and mobile phase B was acetonitrile/isopropanol (60/40; v/v and 4 mM ammonium acetate). The following linear gradient for B was applied: 90 % for 1 min, 90 – 100 % from 1 to 5 min and held at 100 % for 3 min followed by 3 min at initial condition for column re-equilibration. Samples were spiked with cholesterol-d7 (984ng), CE 18:1-d7 (3.291μg) and TG 15:0/18:1-d7/15:0 (528ng) prior to extraction. Triglycerides were analysed in selected ion monitoring (SIM) positive mode, covering a range from TG 32:0 up to TG 56:0 including also unsaturated TGs. MS conditions were: TEM 450°C, GS1 35 psi, GS2 50 psi, CUR 35 psi, IS 5 kV, declustering potential 60 V and entrance potential 10 V. Dwell time was 10 ms. TAGs were quantified using an external calibration with TG 15:0/18:1-d7/15:0. Free cholesterol and CEs were analysed in MRM mode monitoring the precursor to product transitions of 12 CEs and free cholesterol, as [M+NH_4_]^+^. MS conditions were as follows: TEM 150°C, GS1 25 psi, GS2 50 psi, CUR 35 psi, IS 5 kV, declustering potential 70 V, entrance potential 10 V, collision energy 20 V, and collision cell exit potential 25 V. Dwell time was 100 ms for each transition. Cholesterol and CEs were quantified using external calibration curves against the internal standards, with the following primary standards: cholesterol, CE 14:0, CE 16:0, CE 18:0, CE 18:1, CE 20:4 and CE 22:6. For all targeted assays, inclusion criteria for peaks were those at least 5:1 signal-to-noise ratio, and with at least 7 points across the peak.

All targeted lipidomics data were statistically analysed using Students T-test, followed by Benjamini Hochberg correction where any lipid category had >20 variables. All statistical data is provided in Source Data file.

### Untargeted lipidomics

Untargeted lipidomics was conducted using Vero culture Prep 3 (three separate virus samples were extracted, and extracted blanks) on a Waters iClass liquid chromatography system coupled to a Synapt XS QTOF (Waters), in resolution (21,500 FWHM pos, 19,000 FWHM neg) mode. Liquid chromatography was performed at 35 °C using a Waters XBridge Amide column, 3.5 μm, 4.6 × 150 mm, at a flow rate of 0.7 mL/min over 24 min. Mobile phase A was (water/acetonitrile 5/95; v/v and 1 mM ammonium acetate) and mobile phase B was water/acetonitrile (50/50; v/v and 1 mM ammonium acetate). The following linear gradient for B was applied: 0.1 – 6 % B over 6 mins, 6 - 25 % B over 4 mins, 25 - 98 % B over 1 min, 98 – 100 % B over 2 mins. At 13.5 mins, the flow rate changes to 1.5 ml/min and remains at 100 % B until 18.7 min where it returns to 0.1 % B. Flow rate then returns to 0.7 ml/min at 23.5 min. MS conditions were as follows for analysis in negative ion mode: Capillary voltage 1.2kV, source temp 120° C, sampling cone 25, desolvation temp 450 °C, cone gas flow 20, mass range 50 - 2000 amu, scan rate 0.5 sec. Lock mass was Leucine Enkephalin *m/z* 554.2615. For analysis in positive ion mode: : Capillary voltage 1.5 kV, source temp 100° C, sampling cone 30, desolvation temp 500°C, cone gas flow 30, resolution mode, mass range 50 - 2000 amu, scan time 0.5 sec. Lock mass was Leucine Enkephalin *m/z* 556.2771. Prior to feature analysis the data was processed using the Waters compression tool to reduce the noise, changed to centroid using MassLynx and converted to .MZxml by the MSconvert module in Proteowizard. Feature analysis was carried out using the HPLC/QTOF parameters in XCMS online(*37*). The two resulting feature lists (positive and negative) were further processed using the Python program LipidFinder 2.0 in its default configuration(*36*). This includes: solvent, ion fragments, salt clusters, adducts, isotopes and contaminants as well as lipid stacks removal, and outlier and retention time correction. The putative lipid profiling was done using LMSD on LIPID MAPS with 0.05 Da tolerance, searching for “[M-H]^-^”, “[M+H]^+^”, “[M+Na]^+^”, “[M+NH_4_]^+^”, “[M+OAc]^-^” ions and adducts. Next, all matches with deltaPPM >10 were manually removed. Matches to GL in negative ion data, and matches to FA in positive ion data were removed and reassigned as unknowns, with LMSD identifiers removed. In LipidFinder, to remove baseline noise from blanks, the mean of the blank signals for each ion is subtracted from each lipid sample, where they match by RT and m/z value. Then, every frame that is less than 3 times greater than the solvent mean for every sample is removed. After processing, a manual step was also included where ions that were represented in blank samples at >15% the virus sample were judged to be background and removed. Retention time windows based on standards were estimated as follows: lysoPE/PC 10-11 min, PE/PC 6-7 min, Lyso PI 10-12 min, PG 2-4 min, PI 8-10 min, LPG 4.5-5.5 min, MAG/TAG/DG 1-3 min, SM 9-11 min, Cer 1.5-3 min. Note that many ions listed in unknowns are likely to be in source fragments, that will match LMSD entries, that were moved to unknown since they are outside the expected RT window. Mass accuracy is broadly considered down to 3 decimal places. Note that this is a largely unvalidated dataset and provided for further information mining purposes. Data is in SupplementaryData1.xls

### Identification and quantitation of external facing PE and PS on the surface of SARS-CoV2

Total and external PE and PS were derivatised and analysed using LC/MS/MS as described previously(*41*). Briefly, virus particles were suspended in 0.2 ml PBS and incubated with 20 μl of 20 mM NHS-biotin (total PE/PS) or 86 μl of 11 mM EZ-Link Sulfo-NHS-biotin (external PE/PS) for 10 minutes at room temperature before addition of 72 μl of 250 mM L-Lysine. Volumes were increased to 0.4 ml using PBS. Vials containing 1.5 ml chloroform:methanol (1:2) solvent with 10 ng of internal standards (biotinylated 1,2-dimyristoyl-PE and -PS) were used for lipid extraction. The solvent:sample ratio was 3.75:1 as a modified Bligh/Dyer technique (*41*). Following vortexing and centrifugation (400 g, 5 mins), lipids were recovered in the lower chloroform layer, dried under vacuum and analyzed using LC-MS/MS. Samples were separated on an Ascentis C-18 5 μm 150 mm × 2.1 mm column (Sigma Aldrich, USA) with an isocratic solvent (methanol with 0.2 % w/v ammonium acetate) at a flow rate of 400 μl/min. Products were analysed in MRM mode on a Q-Trap 4000 instrument (Applied Biosystems, UK) by monitoring transitions from the biotinylated precursor mass (Q1 *m/z*) to product ion mass (Q3 *m/z*) in negative ion mode. The area under the curve for the analytes was integrated and normalized to internal standards. The ratio of external to total PE/PS was calculated for each molecular species and expressed as a fraction (%)externalised. MRM transitions monitored are provided in Table 5.

**Table 5.** Multiple reaction monitoring (MRM) transitions and instrument settings for the aPL analysed in negative ion mode.

| Analyte | Mass | Biotinylated mass | m/z [M-H]<sup>-</sup> | Biotinylated MRM | DP (V) | CE (V) | CXP (V) |
| --- | --- | --- | --- | --- | --- | --- | --- |
| PE 14:0_14:0 | 635 | 861 | 860 | 860→227 | -135 | -60 | -13 |
| PS 14:0_14:0 | 679 | 905 | 904 | 904→591 | -150 | -42 | -17 |
| PE 18:0p_20:4 | 751 | 977 | 976 | 976→303 | -160 | -60 | -5 |
| PE 18:0a_20:4 | 767 | 993 | 992 | 992→303 | -170 | -58 | -5 |
| PE 16:0p_20:4 | 723 | 949 | 948 | 948→303 | -160 | -60 | -5 |
| PE 18:0a_18:1 | 745 | 971 | 970 | 970→281 | -170 | -58 | -5 |
| PE 18:1p_20:4 | 749 | 975 | 974 | 974→303 | -160 | -60 | -5 |
| PS 18:0a_18:1 | 789 | 1,015 | 1,014 | 1,014→701 | -140 | -44 | -23 |
| PS 18:1a_18:1 | 787 | 1,013 | 1,012 | 1,012→699 | -150 | -46 | -23 |
| PS 18:0a_20:4 | 811 | 1,037 | 1,036 | 1,036→723 | -145 | -42 | -23 |

### Assessment of coagulation activity by Activated Partial Thromboplastin Time (APTT)

Due to logistics of conducting assays with live SARS-CoV2 virus, we used a classical assay that doesn’t require specialist equipment (*81*). Here, the activity of both intrinsic and extrinsic pathways of coagulation are measured in re-calcified plasma activated on contact with a negatively-charged provided by a glass test tube, and the ability of live virus to modulate coagulation was tested. Purified SARS-CoV2 virus was resuspended in PBS, then 50 μl added to 50 μl of normal pooled human plasma (Alpha Laboratories, CCN-10), in a glass tube. As a negative control, 50 μl of PBS was added to plasma instead of virus. Samples were incubated at 37 °C for 1 minute, then 50 μl pre-warmed 20 mM CaCl_2_ added. Samples were incubated at 37 °C and the time until a visible clot formed was measured by visual inspection using a stopwatch. Clot time is defined as any visual evidence for formation of a gel-like structure, recognising that these can either be strong and stable, or looser. In control samples, the fibrin clot was formed in around 2 min (120 sec).

### Western blot

Purity of gradient purified viruses was assessed by Western blot for spike protein (virus) and actin (cells). Virions were resuspended in NuPAGE LDS sample buffer (Thermo) containing 10 % dithiothreitol (DTT), then samples were loaded onto 14 % Tris-Glycine pre-cast gels (Biorad), and run for 1 h at 20 V. Protein was transferred to nitrocellulose membranes by semi-dry transfer, blocked in blocking buffer (5 % non-fat milk in PBST) for 1 h, then stained with primary antibody for 1 h at room temperature. Membranes were washed, incubated with secondary antibody for 1 h at room temperature, washed, developed with Supersignal West Pico (Thermo), and imaged using a G:Box Chemi XX6 (Syngene). Antibodies were all diluted in blocking buffer, and were rabbit anti-actin (A2066, Sigma-aldrich 1:2,000) and mouse anti-SARS-CoV2 Spike (Clone 1A9, Insight; 1:2,000), as well as anti-mouse HRP or anti-rabbit HRP (GE Healthcare; 1:2,000).

### Clinical study design

A four-arm, randomised controlled trial was conducted to study the effectiveness of anti-microbial mouthwashes in vivo. Multicentre Research Ethics Committee approval was obtained (IRAS285247; https://doi.org/10.1186/ISRCTN25647404) and all procedures adhered to the Declaration of Helsinki. Original sample size was calculated based on reported mouthwash activity against enveloped herpes virus; designed with a >80% power to detect a 2-fold reduction in viral load(*58*). Plans for stopping data collection were established in advance; treatment arms would be dropped at interim analysis for efficacy, if a >2-fold reduction in salivary viral load (doi.org/10.1186/ISRCTN25647404) was observed Vs Normasol^®^. However, interim analysis was not possible as with 52 randomised patients recruited, only 15 samples contained live virus at baseline. 406 patients were screened for eligibility at 3 hospitals during a 6 month period. The majority were deemed ineligible for inclusion (see CONSORT flow diagram). Following this, 78 individuals were randomised to receive a mouthwash. On final analysis, 51/78 patients had no live SARS CoV2 in baseline salivary samples and were excluded (see CONSORT flow diagram). No other patients were excluded and no outlying data removed, leaving 27 patients. The primary and secondary endpoints established prior to the study were: viral load of SARS CoV2 at 30 mins and viral load of SARS CoV2 at 1,15 and 60 mins (https://doi.org/10.1186/ISRCTN25647404). Sample collection was stopped after 6 months due to falling patient numbers and the data were independently analysed.

Briefly, following informed consent, in-patients with PCR-confirmed COVID-19 infection within the last 14 days, were recruited at the University Hospital of Wales, the Royal Glamorgan Hospital and Betsi Cadwalader University Health Board in Wales UK. Participants were assigned to one of the four arms: Dentyl Dual Action (CPC); Videne (povidone-iodine); SCD Ultra (CPC) or Normasol^®^ (sterile saline 0.9% [w/v]) using a balanced randomisation scheme (provided by Dr Damian Farnell, Cardiff University). Baseline saliva was collected into 30 mL Universal containers. The patient then rinsed their mouth for 30 seconds with 10 mL of the mouthwash. Saliva samples were then collected after 1, 15, 30 and 60 minutes into sterile 30 mL Universal containers. Anonymised samples were transported and stored at -80°C and transferred to the approved BSL3 facility at Cardiff University where live virus was titrated as above. Results were expressed as log_2_ fold change from baseline. Clinical and research staff involved in sample collection and laboratory analysis were blinded as to which product was which.

### Statistical analysis for clinical study

At termination of the study, blinded data was analysed by an independent observer (RGN) who was not involved in the design of the clinical trial or randomisation. All original data on viral load (PFU/mL of saliva), together with the Log_2_ fold change in from baseline to 60 minutes post-treatment is presented. The viral load from baseline to 60 minutes post-treatment was used to calculate a geometric mean ratio (mean change from baseline to post-treatment). Data was then log transformed. Where virus was not detected, zero values were replaced with 4, just below the lower limit of detection for the assay (5 PFU/ml). Lower and upper confidence limits from the relevant mean and SD on the transformed scale and t and p-values were determined using unpaired t test (Table 5).

## Data Availability

All data is included in the manuscript

## Author contributions

MBP, VJT, DW, PDSR, JAJ, AZ conducted lipidomics analysis, VOD, RCM, HK, WJG reviewed lipidomics data, DWT, MP and AA designed the clinical study, AA, DWT, JH, RM, DO, CAL supervised the clinical recruitment and sample-collection in individual Heath Boards. NB prepared and provided surfactant solutions, and designed experiments. VOD, RJS, DWT designed the study, supervised the data collection and analysis and drafted the manuscript. ZS, ES, AR, KB, RJS performed all *in vitro* analysis and preparation of samples. RGN undertook independent statistical analysis. All authors edited and approved the manuscript. We acknowledge intellectual input from Matthew Conroy, LIPID MAPS Biocurator, technical support from Valeria Grossini, and Professor Adrian Maunder for his help in clinical study design.

## Figure Legends

**Supplementary Figure 1.**
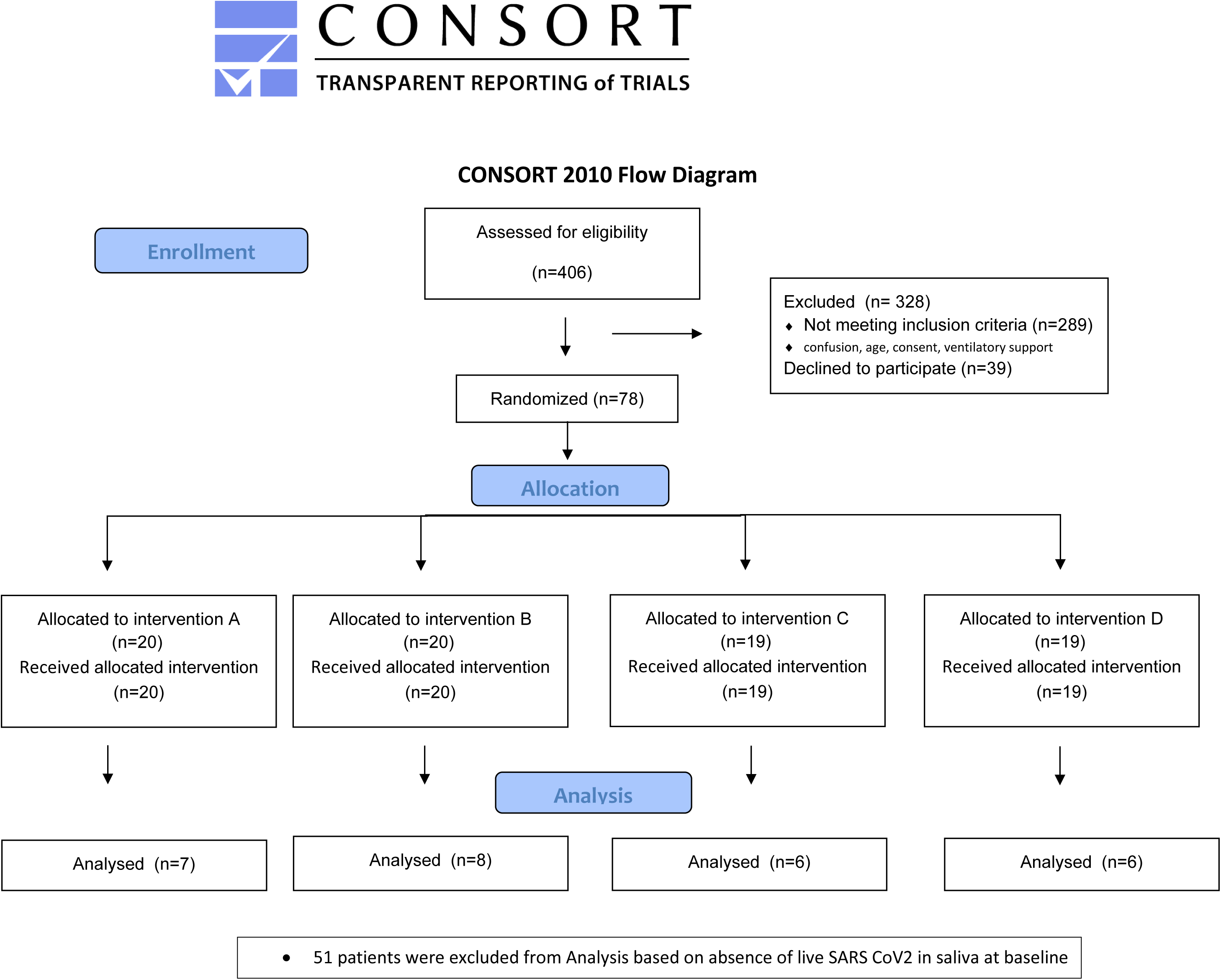
Uncropped western blots, relating to Figure 1B. Boxed lanes relate to the figure shown, other lanes relate to a different study and are not relevant. Lane 3 = purified virus, Lane 4 = mock infected cells, Lane 5 = infected cells. *Panel A. Immunostaining of spike protein*. The membrane was stained with mouse anti-SARS-CoV2 Spike antibody and exposed for 30 sec. Bands corresponding to S1/S2 (∼200kDa), and cleaved S1 (∼80kDa) domains are observed. *Panel B. Immunostaining of actin*. The gel was stripped and reprobed with rabbit anti-actin antibody, then exposed for 3 min. Note that in addition to actin (∼40kDa), incomplete stripping of anti-Spike antibody was also observed.

